# A Whole-Blood Four-Gene Panel Distinguishes Pediatric Crohn’s Disease from Disorders of Gut-Brain Interaction

**DOI:** 10.64898/2026.11.16.26367283

**Authors:** Cameron Lunn, Kara Margolis, Pinar Kiper Misirlioglu, Madeline Xin, Vasuda Kapoor, Yelena Afanasyeva, Lindsey Albenberg, David Piccoli, Janet Okraku-Mantey, Paul Zapille, Peter Meyn, David Fenyö, Jeremiah Levine, Leonardo Trasande

## Abstract

**Background and Aims:** Crohn’s disease and disorders of gut-brain interaction present with overlapping symptoms, and distinguishing them often requires endoscopy. We assessed whether the whole-blood transcriptome at presentation distinguishes these conditions and reflects disease activity.

**Methods:** In a prospective two-center cohort, whole-blood RNA sequencing was performed in 425 treatment-naïve participants aged 5 to 25 years (112 Crohn’s disease, 313 disorders of gut-brain interaction). Differential expression was modeled for diagnosis and, within Crohn’s disease, for the Pediatric Crohn’s Disease Activity Index. Co-expression modules were integrated with protein–protein interaction networks, and machine learning distilled network genes into a diagnostic panel, evaluated internally and in an independent pediatric cohort.

**Results:** Of 1,095 diagnosis-associated genes, 377 were also associated with disease activity, all with concordant direction. Five co-expression networks were perturbed, spanning neutrophil degranulation, hemostasis, interferon signaling, and T-cell and cytotoxic lymphocyte programs. A 4-gene panel (*PSTPIP2*, *CLIC4*, *CACNB4*, *GZMK*) drawn from 4 networks achieved an area under the receiver operating characteristic curve of up to 0.96 in held-out samples and up to 0.93 externally, with external specificity and positive predictive value of up to 0.95 and 0.93, respectively. The panel matched or exceeded every single-network panel with fewer genes, and minimally active Crohn’s disease overlapped transcriptionally with disorders of gut-brain interaction.

**Conclusions:** At diagnosis, Crohn’s disease is distinguished from disorders of gut-brain interaction by a systemic signature of neutrophil and platelet activation and lymphocyte depletion that scales with disease activity. A 4-gene whole-blood panel may help prioritize symptomatic children for endoscopy and warrants prospective validation.

## INTRODUCTION

Crohn’s disease (CD) is a chronic inflammatory disorder of the gastrointestinal (GI) tract with increasing prevalence and significant financial burden worldwide (1). CD is transmural across the GI tract, causing manifestations that vary from mild abdominal pain and inflammation to severe bowel injury and/or obstruction, occasionally leading to surgical resection (2). Disorders of gut-brain interaction (DGBI), including irritable bowel syndrome (IBS) and functional dyspepsia, are functional gastrointestinal disorders marked by recurrent pain, altered bowel habits, bloating, and/or dyspeptic symptoms, symptoms that can mimic those of active inflammatory bowel disease (IBD) (3, 4). Among children with IBD in remission, IBS and DGBI with abdominal pain occur in up to 16.1% and 29.5%, respectively, further complicating the differentiation between a diagnosis of CD and DGBI (5). Due to these challenges, diagnosis requires extensive non-invasive and invasive testing (6), including endoscopy, histology, and imaging (6–8). Still, these diagnostic methods are not 100% reliable (9).

Currently, there is no cure for CD, so clinical management aims to minimize severity by inducing and maintaining disease remission (10). A widely utilized method to track pediatric CD disease activity clinically is the Pediatric Crohn’s Disease Activity Index (PCDAI), which ranges from 0 to 100 based on a combination of questionnaire responses and biological measurements, enabling severity-aware assessment in practice and research (11, 12). Many of the questions within the PCDAI, however, are also relevant to DGBIs, further complicating the diagnoses.

CD pathogenesis is generally characterized by dysregulated innate and adaptive immunity, epithelial barrier dysfunction, intestinal microbiome alterations, and genetic risk factors (13). Bulk RNA sequencing (RNA-seq) CD studies suggest prominent roles for neutrophil activation/degranulation and innate-immune signaling pathways including NOD-like and Fcγ-receptor-driven pathways, alongside Th1/Th17 cell responses (14), all of which are associated with disease activity (15). Several of these pathways, including TNF- and IL-12/23-driven inflammation, are targets for current therapies, yet non-response and loss of response remain common in clinical practice (16, 17). These features are thus driving efforts to develop biomarkers that can distinguish CD from DGBI, as well as aid clinicians in facilitating earlier diagnosis and treatment stratification. However, most studies utilize tissue biopsy samples (18–21). Whole blood offers a less-invasive alternative that is obtainable at presentation, captures systemic immune activity, and is amenable to scalable clinical testing. Most whole-blood studies, however, are limited to non-symptomatic healthy controls and/or assess disease activity at time of surgery (22–25). There are few studies investigating whole-blood gene expression at (a) time of CD diagnosis and (b) utilizing symptomatic controls (26, 27). Because many individuals with suspected IBD initially present with functional gastrointestinal symptoms, we used DGBI as a symptomatic, non-IBD comparison population to minimize bias inherent to healthy control designs.

We therefore performed whole-blood RNA sequencing in a prospective, two-center cohort of treatment-naïve children and young adults presenting with undiagnosed gastrointestinal symptoms, comparing those ultimately diagnosed with CD with those diagnosed with DGBI. To our knowledge, this is the largest transcriptome-wide whole-blood study of treatment-naïve pediatric CD and the first pediatric study to use a Rome IV-defined DGBI comparator. Because patients present across a range of disease activities, we also modeled PCDAI within CD. This allowed us to ask whether the signal that separates CD from DGBI is the same signal that tracks clinical severity. Our aims were to (1) define transcriptional and immune cell-composition differences between CD and DGBI and their relationship to PCDAI; (2) organize these differences into co-expression and protein–protein interaction networks to characterize the biological programs involved at a systems-level; and (3) identify, within each network, the genes that most reproducibly discriminated CD from DGBI, combine them into a minimal cross-module diagnostic panel, and evaluate its predictive performance in an independent pediatric cohort composed of CD and symptomatic control whole-blood samples.

## MATERIALS AND METHODS

Complete methods are provided in the Supplementary Materials.

### Study design, sample collection, and ethical approval

All participants (and guardians, where appropriate) provided written informed consent, and study procedures were approved by the Institutional Review Boards of the NYU Grossman School of Medicine (i19-01024) and the Children’s Hospital of Philadelphia (22–020141). This study drew on participants from a prospective observational cohort. Between September 2019 and June 2024, individuals aged 5–25 years presenting with undiagnosed gastrointestinal symptoms at NYU Langone Health, New York, NY (NYU) or the Children’s Hospital of Philadelphia, Philadelphia, PA (CHOP) were enrolled. Patients were ultimately diagnosed with either (a) CD by standard radiologic and histologic criteria or (b) DGBI according to the Rome IV criteria (28). Individuals with biopsy-confirmed ulcerative colitis, celiac disease, or CD who received treatment prior to sample collection were excluded. The site study team independently reviewed all CD diagnoses using established criteria that incorporated clinical history, physical examination, laboratory testing, and endoscopic and/or radiologic evaluation. Disease activity in CD was assessed with the PCDAI, and patients were stratified into minimally-active (PCDAI ≤ 10), mild (10 < PCDAI ≤ 30), or moderate-severe (30 < PCDAI) categories based on accepted thresholds from previous studies (29, 30). We used the term minimally-active instead of remission because individuals presenting to the clinic for the first time with gastrointestinal symptoms cannot have achieved remission. These groups were used to summarize the CD population and visualize results but were not used as model inputs.

Across the study period, 438 peripheral blood samples were collected during (a) first presentation to the clinic or (b) follow-up for the diagnostic tests. Thirteen samples were excluded at quality control, as described below, yielding a final analytic cohort of 425 samples (112 CD, 313 DGBI) used for all downstream analyses. C-reactive protein (CRP) was measured in a subset of our participants, and demographic data were collected by the study team, which included age, BMI percentile, secondhand smoke exposure, and sex. Missing secondhand smoke exposure (n = 31) was coded as a separate category in downstream models. Demographic and clinical characteristics were summarized by study group and compared using Wilcoxon rank-sum tests for continuous variables and Pearson chi-square tests for categorical variables.

### Whole-Blood RNA-seq

Total RNA was extracted from whole blood, and quality was verified by Bioanalyzer trace. Library preparation was performed using the Tecan RNA library preparation kit. The protocol included ribosomal RNA (rRNA) depletion and integrated DNase treatment. For each sample, 5 ng of total RNA was used as input, and 10 PCR cycles were applied during library amplification. Library concentrations were quantified by quantitative PCR, and samples were pooled to achieve uniform sequencing depth. Sequencing was performed on the Illumina NovaSeq X Plus platform, generating 2 × 51 bp paired-end reads, with one plate loaded per flow cell.

### Bioinformatic Analyses

Bulk RNA-seq reads were processed with a standard nf-core/Nextflow pipeline (doi:10.5281/zenodo.1400710) (31), which included STAR alignment to GRCh38, and transcript quantification with salmon. Thirteen samples with STAR alignment < 85% were removed. Quantification files were imported into R v4.4.1 (32), where transcripts were summarized to the gene level, and low-expression genes were removed with edgeR. Batch effects were investigated using principal component analysis (PCA). Bulk deconvolution (CIBERSORT/LM220) (33) was performed to estimate immune cell fractions between CD and DGBI groups, where the trimmed mean of M-values (TMM) normalized counts per million (CPM) matrix was used as input. Estimated immune cell fractions were compared between CD and DGBI with Wilcoxon tests. Differential expression analysis (DEA) was performed with the limma-voom pipeline to identify differentially expressed genes (DEGs). We fit two models: (a) modeling diagnosis (CD-vs-DGBI) using all samples; and (b) modeling continuous PCDAI using only the CD samples. DEGs in the diagnosis model represent genes differentially expressed between CD and DGBI samples (false discovery rate [FDR] < 0.05; | log_2_ fold change | [log_2_FC] ≥ 0.60); while the PCDAI model identifies DEGs whose expression is associated with worsening CD clinical severity (FDR < 0.05, | log_2_FC | ≥ 0.02; log_2_FC is estimated per PCDAI point). Both models were adjusted for age, sex, sequencing batch, recruitment site, and secondhand smoke exposure. To characterize shared pathway-level perturbations across both CD activity and diagnosis, we took the overlapping DEGs from both models and performed over-representation analysis using Gene Ontology Biological Process (GOBP) and Kyoto Encyclopedia of Genes and Genomes (KEGG).

Weighted Gene Co-Expression Network Analysis (WGCNA) was performed using the normalized residual counts (batch-corrected) to identify co-expressed gene modules (34). The expression profile of each module was summarized by its module eigengene (ME), the first principal component of the module’s gene expression, which was then correlated with CD diagnosis and PCDAI group variables. Module genes were then overlapped with DEGs from both models to identify differentially co-expressed modules (DCMs) enriched for DEGs associated with CD diagnosis and CD severity. The DCMs’ gene sets were further refined by filtering the modules into respective protein-protein interaction (PPI) networks with Search Tool for the Retrieval of Interacting Genes/Proteins (STRING v12.0). Combined score ≥ 500 was used to filter for interactions between protein-coding genes. Each differentially expressed PPI network was pruned to the largest connected component (LCC) and summarized by proportion of network nodes retained, the global clustering coefficient, and Louvain modularity. Reactome pathway over-representation analysis was performed separately for each module’s LCC using all genes tested from the two models as the background, with significance defined as FDR < 0.05. Nodes were annotated by their DEA results from both models (log_2_FC and FDR significance), and we defined a composite centrality as the mean rank across unweighted degree, betweenness centrality, PageRank centrality, and k-core. We used the composite centrality and DEA results to identify differentially expressed hub genes. Community structure within each module’s LCC was inferred with a nested stochastic block model (NSBM), fit by minimizing the model’s description length. The resulting finest-level partition was used for network visualization and to test block-level concentration (clusters) of DEGs.

### Supervised Machine Learning

DEGs within each DCM’s LCC were the candidate features. Samples were split 70/30 into training and held-out test partitions, stratified by diagnosis and CD activity group. In Stage 1, each module was analyzed on the training partition by repeated nested cross-validation (CV; 10 folds, 10 repeats), with least absolute shrinkage and selection operator (LASSO) gene selection followed by LASSO, random forest (RF) and support vector machine (SVM) classifiers. Genes selected in at least 90% of 1,000 bootstrap LASSO fits formed each module’s stable genes. In Stage 2, stable genes were pooled across modules, and genes selected in at least 85% of a second round of bootstraps formed the final panel. Because the panel genes were fixed before CV, the final-panel CV was not nested with respect to gene selection. The panel was evaluated once on the held-out test partition and, after refitting on all 425 samples, once on 39 CD and 37 symptomatic control (SC) whole-blood samples from GSE126124 (26), with expression z-scored within each cohort. This external test cohort included participants aged 8 to 18 years, and the SC population consisted of individuals with juvenile polyps, infectious colitis, and functional intestinal disorders (two controls who were later diagnosed with CD were excluded). Thresholds were Youden-optimal cutoffs from the internal out-of-fold predictions. We report area under the receiver operating characteristic curve (AUC-ROC) with 95% CIs alongside area under the precision-recall curve (AUC-PR), sensitivity, specificity, positive and negative predictive value (PPV, NPV), F1 score, balanced accuracy, and Brier score. Single-gene, leave-one-out, and subgroup misclassification analyses, and all tuning details are described in the Supplementary Materials.

## RESULTS

### Study participants

In total, 425 high-quality whole-blood samples were collected from untreated participants in our study, which comprised 112 CD and 313 DGBI participants (Table 1). The median age was 13 years, and 78% of our population was enrolled from NYU and 22% from CHOP. The median PCDAI in the CD group was 28, with 11 individuals classified as minimally-active, 59 as mild, and 42 as moderate-severe. The single largest subclassification in our DGBI group was IBS, which comprised 34% of the DGBI samples. Functional abdominal pain-not otherwise specified, functional dyspepsia, and functional constipation were the next most common classifications, comprising 28%, 18%, and 14%, respectively. Functional nausea and vomiting, nonretentive fecal incontinence, abdominal migraine, aerophagia, and rumination syndrome represented less than 6% of our DGBI population. CD participants were more often male than DGBI participants (P = 0.017), whereas secondhand smoke exposure did not differ across CD and DGBI groups. Both CRP (mg/L) and BMI percentiles were significantly different across groups, with medians of 13 vs 0 mg/L for CRP and 30th vs 58th percentile for BMI for CD and DGBI groups respectively (both P < 0.001). CRP was unable to be measured in 102 of our participants.

**Table 1.** Characteristics of study participants.

| Characteristic | Overall<br>N = 425 | CD<br>n = 112 | DGBI<br>n = 313 | P value |
|---|---|---|---|---|
| Age, years, median (IQR) | 13.0 (10.0, 16.0) | 13.0 (10.1, 15.0) | 13.0 (10.0, 16.0) |  |
| Sex, n (%) |  |  |  | 0.017 |
| Female | 210 (49%) | 44 (39%) | 166 (53%) |  |
| Male | 215 (51%) | 68 (61%) | 147 (47%) |  |
| Recruitment site, n (%) |  |  |  |  |
| CHOP | 94 (22%) | 25 (22%) | 69 (22%) |  |
| NYU | 331 (78%) | 87 (78%) | 244 (78%) |  |
| PCDAI, median (IQR) | – | 28 (18, 36) | – |  |
| PCDAI group, n (%) |  |  |  |  |
| Minimally active | – | 11 (9.8%) | – |  |
| Mild | – | 59 (53%) | – |  |
| Moderate-severe | – | 42 (38%) | – |  |
| DGBI subtype, n (%) |  |  |  |  |
| IBS | – | – | 107 (34%) |  |
| FAP-NOS | – | – | 88 (28%) |  |
| Functional dyspepsia | – | – | 56 (18%) |  |
| Functional constipation | – | – | 44 (14%) |  |
| Functional nausea and vomiting | – | – | 12 (3.8%) |  |
| Nonretentive fecal incontinence | – | – | 3 (1.0%) |  |
| Abdominal migraine | – | – | 1 (0.3%) |  |
| Aerophagia | – | – | 1 (0.3%) |  |
| Rumination syndrome | – | – | 1 (0.3%) |  |
| Secondhand smoke exposure<sup>b</sup>, n (%) |  |  |  |  |
| No | 351 (89%) | 99 (89%) | 252 (89%) |  |
| Yes | 43 (11%) | 12 (11%) | 31 (11%) |  |
| BMI percentile, median (IQR) | 50 (21, 78) | 30 (6, 61) | 58 (28, 82) | <0.001 |
| CRP, mg/L, median (IQR)<sup>a</sup> | 1 (0, 6) | 13 (4, 47) | 0 (0, 2) | <0.001 |

Values are n (%) or median (interquartile range). P values from Wilcoxon rank-sum tests (continuous variables) or Pearson chi-square tests (categorical variables). Percentages for PCDAI and DGBI subgroups are of the CD and DGBI groups, respectively.
<sup>a</sup>CRP could not be measured in 11 CD and 91 DGBI participants. <sup>b</sup>Secondhand smoke exposure was missing for 31 participants (1 CD, 30 DGBI).
Abbreviations: BMI, body mass index; CD, Crohn’s disease; CHOP, Children’s Hospital of Philadelphia; CRP, C-reactive protein; DGBI, disorders of gut-brain interaction; FAP-NOS, functional abdominal pain–not otherwise specified; IBS, irritable bowel syndrome; IQR, interquartile range; NYU, NYU Langone Health; PCDAI, Pediatric Crohn’s Disease Activity Index.

### Whole-blood gene expression profiles separate CD from DGBI and capture within-CD disease activity

To assess transcriptomic differences between children newly diagnosed with CD and DGBI, we performed RNA sequencing of 425 high-quality whole-blood samples. The median library size was 16.9 million reads and the median STAR alignment rate was 89%. Pseudogenes, hemoglobin genes, and low-signal genes were filtered out, yielding 15,444 highly expressed genes for downstream analysis. Batch effects from covariates were assessed using PCA, where sequencing batch was the largest contributor of unwanted variance (Supplementary Figure S1). We regressed out the covariate effects of sequencing batch, sex, secondhand smoke, recruitment site, and age and re-ran PCA on the batch-corrected normalized matrix. The batch-associated separation, specifically the sequencing batch split, was eliminated (Figure 1A). Separation between CD and DGBI samples increased following correction. PC1 and PC2 accounted for 53.4% and 16.1% of the variance, respectively, and revealed two distinct clusters primarily defined by diagnosis of CD (purple) or DGBI (orange). More specifically, the moderate-severe samples (pluses) clustered distinctly from DGBI samples (circles), whereas minimally-active CD samples (triangles) mostly overlapped with the DGBI group. Likewise, the mild CD samples (squares) tended to fall between these clusters. Altogether, this suggested a dual gradient between DGBI and CD samples, as well as from minimally-active CD to moderate-severe CD states.

**Figure 1.**
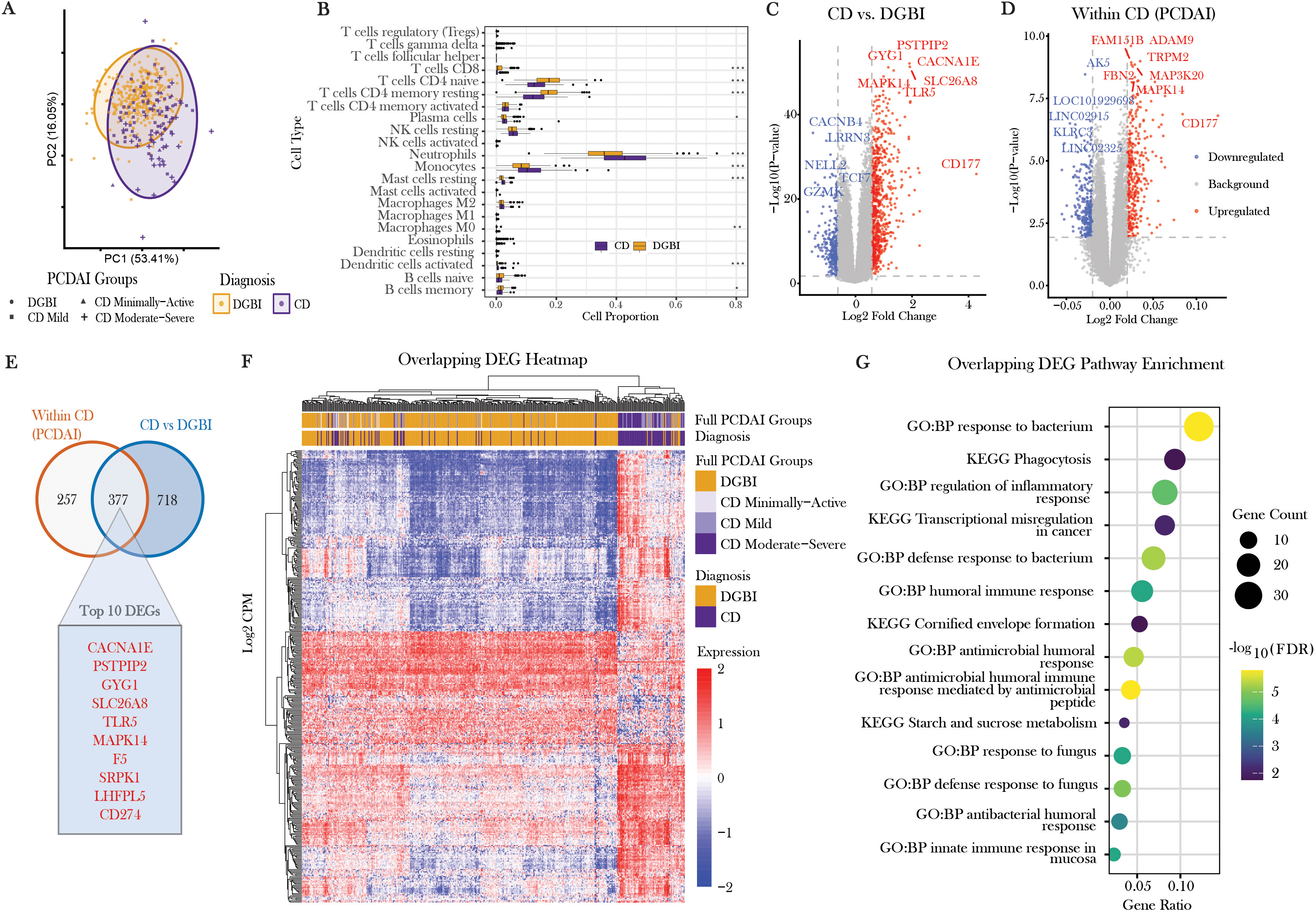
Whole-blood transcriptomic differences between CD and DGBI, and in relation to CD activity. (A) Principal component analysis of whole-blood gene expression, plotting PC1 against PC2. Point shape indicates PCDAI group (DGBI are circles; minimally-active CD are triangles; mild CD are squares; and moderate-severe CD are plus signs), and color indicates diagnosis (DGBI is orange; CD is purple), with ellipses drawn for each diagnosis. (B) Box plots of estimated immune cell proportions for each cell type in CD (purple) and DGBI (orange). (C) Volcano plot of differential expression between CD and DGBI. (D) Volcano plot of differential expression in relation to PCDAI within-CD. In both volcano plots, the x-axis shows log_2_FC, the y-axis shows −log_10_(P value), and dashed lines mark the significance and fold-change thresholds. Upregulated genes are shown in red, downregulated genes in blue, and nonsignificant genes in gray, with top genes labeled. (E) Venn diagram of DEGs identified within-CD by PCDAI (n = 257 unique), CD-vs-DGBI (n = 718 unique), and in both models (n = 377). The inset lists the top 10 DEGs shared by both analyses. (F) Heatmap of scaled expression (range, −2 to 2) of the DEGs shared by both analyses (rows) across samples (columns), with hierarchical clustering dendrograms. Annotation bars indicate PCDAI group and diagnosis. (G) Dot plot of GO biological process and KEGG pathway terms enriched among the shared DEGs. The x-axis shows gene ratio, dot size indicates the number of genes, and color indicates - log_10_(FDR). Abbreviations: CD, Crohn’s disease; DEG, differentially expressed gene; DGBI, disorders of gut-brain interaction; FDR, false discovery rate; GO, Gene Ontology; KEGG, Kyoto Encyclopedia of Genes and Genomes; log_2_FC, log_2_ fold change; PC, principal component; PCDAI, Pediatric Crohn’s Disease Activity Index. **Alt text:** Graphs labeled A to G. A is a scatterplot of principal component 1 versus principal component 2, with points colored orange for DGBI and purple for CD, point shapes indicating PCDAI group, and an ellipse drawn around each diagnosis. B is a set of boxplots comparing estimated proportions of 22 immune cell types between CD and DGBI. C is a volcano plot of log_2_ fold change versus −log_10_ P value for CD versus DGBI, with upregulated genes in red, and downregulated genes in blue, nonsignificant genes in gray, and top genes labeled. D is a volcano plot in the same format for genes associated with PCDAI within CD samples. E is a Venn diagram showing 718 DEGs unique to CD versus DGBI model, 257 unique to the PCDAI model, and 377 shared, with top 10 of the shared genes shown. F is a heatmap of scaled expression of the shared genes across all samples, with hierarchical clustering dendrograms and annotation bars for diagnosis and PCDAI groups. G is a dotplot of enriched GO and KEGG pathways, with pathways on the y-axis and gene ratio on the x-axis, dot size showing gene count, and a color gradient showing significance.

### Immune cell deconvolution and differential gene expression analyses reveal broad inflammatory program separating CD and DGBI samples and that intensifies with CD severity

To assess whether leukocyte composition differed between CD and DGBI samples, we utilized CIBERSORT to estimate the proportions of 22 immune-cell types. As shown in Figure 1B, several cell populations differed significantly between the groups based on a Wilcoxon test with an FDR < 0.05. CD samples exhibited higher proportions of monocytes, neutrophils, activated mast cells, plasma cells, activated dendritic cells, and M0 macrophages compared with DGBI samples, but lower proportions of CD8+ T cells, naïve CD4+ T cells, resting memory CD4+ T cells, activated memory CD4+ T cells, and both naïve and memory B cells compared with DGBI.

To further investigate transcriptional differences between CD and DGBI samples, we implemented two DEA models. The first comparison between CD and DGBI samples identified 1,095 DEGs, with 757 up-regulated and 338 down-regulated (Figure 1C, Supplementary Table S1). Prominently up-regulated genes in CD samples compared to DGBI samples included *PSTPIP2*, *CD177*, *GYG1*, *TLR5*, *SLC26A8*, *CACNA1E*, and *MAPK14*. The top downregulated genes in CD included *CACNB4*, *LRRN3*, *NELL2*, *GZMK*, and *TCF7*. The PCDAI model using only CD samples identified 634 total DEGs, with 365 upregulated and 269 downregulated (Figure 1D, Supplementary Table S2). PCDAI-associated upregulated DEGs included *CD177*, *MAPK14*, *MAP3K20*, *ADAM9*, and *TRPM2*, with down-regulated genes *KLRC3*, *LOC101929698*, *LINC02915*, and *AK5*. Several genes like *MAPK14* and *CD177* were ranked in the top 10 of both models, which motivated an investigation into their overlapping signature.

Comparing DEG sets of CD-vs-DGBI and within-CD differential expression models revealed 377 overlapping genes, while 718 and 257 genes were unique to the CD diagnosis and within-CD models, respectively (Figure 1E). All 377 overlapping DEGs exhibited directionally concordant gene coefficient effect sizes, indicating that the molecular signatures distinguishing CD from DGBI intensified in alignment with CD severity (Supplementary Table S3). A hierarchical clustering heatmap of the 377 overlapping DEGs (Figure 1F) showed clear CD/DGBI separation with gradients that aligned to PCDAI groups. Like the PCA plot, most CD samples that were clustered with the DGBI samples were minimally-active and mild participants, whereas the divergent cluster of CD samples was largely comprised of the moderate-severe and mild samples. Pathway enrichment using KEGG and GOBP of the overlapping DEGs (Figure 1G) highlighted inflammatory programs and response to microbes. Specifically, response to bacterium, phagocytosis, regulation of inflammatory process, Transcriptional misregulation in cancer, humoral immune response, Cornified envelope formation, Starch and sucrose metabolism, response to fungus, and innate immune response in mucosa were the top unique pathways enriched (full list of enriched pathways is in Supplementary Table S4). Our results suggest that the antimicrobial and immune system responses underlying disease activity in CD are also shared with those that differentiate CD from DGBI.

### WGCNA identifies CD-associated co-expression modules

To contextualize the results from our differential expression models, we assessed the co-expression programs related to CD diagnosis and PCDAI severity by constructing a signed co-expression network with WGCNA. The scale-free topology fit increased monotonically and exceeded 0.80 for soft-threshold power = 8, while mean connectivity declined as expected (Supplementary Figure S2A, S2B). Thus, power = 8 was used for WGCNA module construction, which identified a total of 16 modules, plus the unassigned grey module. The eigengene dendrogram/heatmap showed blocks of strongly co-varying modules (Supplementary Figure S2C). Positively co-regulated modules clustered together, while anticorrelated modules segregated to opposing branches, suggesting distinct biological groupings. Module sizes spanned a broad range (Supplementary Figure S2D). The unassigned grey module contained 3,400 genes. Among colored modules, turquoise (2,645) was largest, followed by blue (2,287), brown (1,603), yellow (1,510), green (1,117), red (901), and black (688). Smaller modules comprised pink (224), magenta (184), purple (174), greenyellow (145), tan (144), salmon (133), cyan (127), midnightblue (83), and lightcyan (79).

### Differential module expression analysis and integration with differentially expressed genes

To assess how the co-expression modules related to disease diagnosis and severity, we correlated MEs with DGBI, minimally-active CD, mild CD, or moderate-severe CD groupings (Figure 2A). The most significant positive correlations for diagnosis were blue (r = 0.48, P < 0.001), black (r = 0.36, P < 0.001), cyan (r = 0.36, P < 0.001), and red (r = 0.30, P < 0.001). Weaker positive correlations (r < 0.30) were observed in the midnightblue (r = 0.24, P < 0.001) and green (r = 0.14, P < 0.001) modules. Positive correlation indicated that each module’s respective gene sets are generally upregulated in CD-vs-DGBI samples. The most significant negative associations were turquoise (r = −0.52, P < 0.001), salmon (r = −0.37, P < 0.001), tan (r = −0.32, P < 0.001), and greenyellow (r = −0.30, P < 0.001), indicating the genes in these modules were downregulated as a whole in CD versus DGBI. Weaker negative correlations were observed in the magenta (r = −0.29, P < 0.001), brown (r = −0.29, P < 0.001), yellow (r = −0.24, P < 0.001), and lightcyan (r = −0.24, P < 0.001) modules. Accordingly, the strength of module-trait correlations decreased progressively from the moderate-severe CD subgroup to the mild and minimally-active CD subgroups, reflecting a gradient in transcriptional association of each co-expression module’s set of genes with CD disease severity (Supplementary Figure S2E).

**Figure 2.**
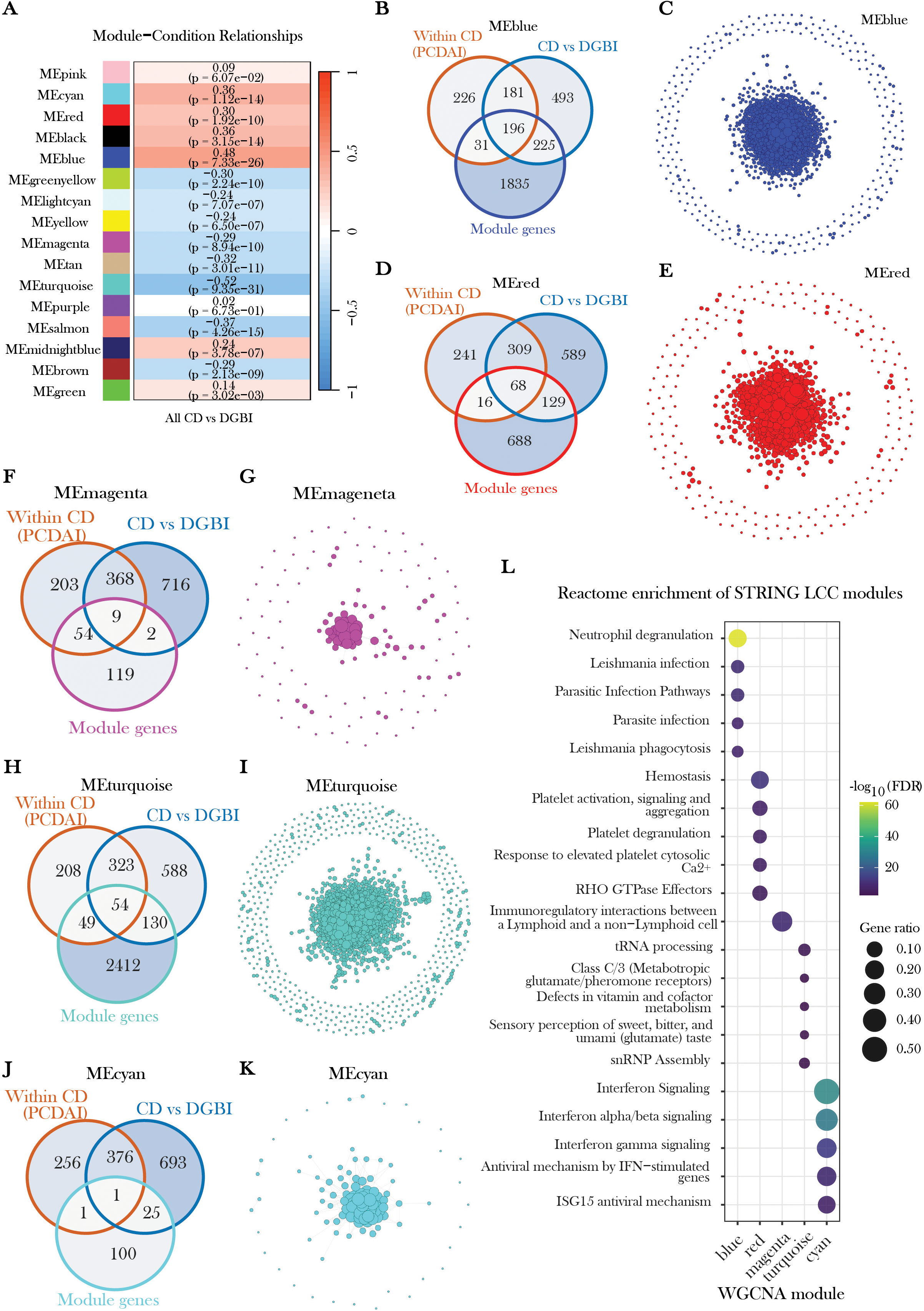
Weighted gene co-expression network analysis identifies five differentially co-expressed modules (DCMs) encompassing a spectrum of biological pathways, ranging from neutrophil degranulation to interferon signaling. (A) Module-trait correlation analysis identified 6 modules positively correlated and 8 modules negatively correlated with Crohn’s disease (CD) diagnosis, while 2 modules were uncorrelated. The greatest overlap with differentially expressed genes from the CD diagnosis and CD activity (Pediatric Crohn’s Disease Activity Index [PCDAI]) models was observed in the (B) blue, (D) red, (F) magenta, (H) turquoise, and (J) cyan modules. Protein-protein interaction networks reconstructed using the Search Tool for the Retrieval of Interacting Genes/Proteins (STRING) for the (C) blue, (E) red, (G) magenta, (I) turquoise, and (K) cyan DCMs identified a core, largest connected component (LCC) used for downstream pathway and network analyses. Smaller components and singletons are included in each plot. (L) The top five pathways identified by Reactome overrepresentation analysis demonstrated that the blue LCC was enriched for neutrophil degranulation and parasitic infection pathways; the red LCC was characterized by hemostasis and platelet activation; the smaller magenta LCC formed a network enriched solely for immunoregulatory interactions between lymphoid and non-lymphoid cells; the turquoise LCC was enriched for tRNA processing and small nuclear ribonucleoprotein (snRNP) assembly; and the denser cyan LCC was enriched for interferon and antiviral signaling. Abbreviations: DCM, differentially co-expressed module; LCC, largest connected component; STRING, Search Tool for the Retrieval of Interacting Genes/Proteins. **Alt text:** Graphs and networks labeled A to L. A is a heatmap of correlations between module eigengenes and CD diagnosis and activity groups, with a color scale gradient showing correlation. B, D, F, H, and J are Venn diagrams showing overlap between the module genes and differentially expressed genes for the blue, red, magenta, turquoise, and cyan modules. C, E, G, I, and K are protein-protein interaction networks for each module, with the largest connected component in the middle, surrounded by singleton and smaller components. L is a dot plot of the top 5 Reactome pathways for each module, with pathways on the y-axis and color gradient showing significance.

To directly evaluate DCMs, we assessed the overlap of our DEGs from our differential expression results with each module’s gene sets and recorded the proportion of DEGs in each module. We observed the highest overlap in the blue, red, magenta, turquoise, and cyan modules, which we deemed the DCMs (Figure 2B, 2D, 2F, 2H, 2J respectively). The greenyellow, purple, black, brown, green, salmon, yellow, tan, pink, and lightcyan modules had less than 5% overlap with the CD-vs-DGBI and within CD DEG sets, so these remaining modules were discarded from downstream interaction analyses. Additionally, the midnightblue module was dropped from downstream network analysis as it was predominantly comprised of immunoglobulin variable-region genes not represented in the STRING database.

### PPI network analysis of differentially co-expressed modules

To resolve our 5 DCMs into interaction networks, we mapped the genes in each module to STRING and retained high-confidence interactions to construct module-specific PPI networks. Mapping rates ranged from ∼80% to 95% across modules since STRING only contains protein-coding gene interactions (Supplementary Table S5).

Across the 5 DCM networks, the LCC varied in size, from 39% of nodes in magenta to 87% in blue, with the remainder of nodes forming singletons and small disconnected components (Supplementary Table S5). Each LCC nonetheless contained >98% of the edges in its full network, indicating that each module’s interactions were concentrated within a single core component (Figure 2C, 2E, 2G, 2I, 2K). Because the LCC captures this densely interacting core, we restricted downstream analysis to the LCC and dropped disconnected nodes. LCC topology differed across modules (Supplementary Table S5). Clustering coefficients were highest in cyan (0.75) and magenta (0.66) and lowest in blue (0.24) and turquoise (0.36), while red exhibited moderate clustering (0.52). Modularity showed the inverse pattern, highest in turquoise (0.69), red (0.63), and blue (0.50) and lowest in cyan (0.11) and magenta (0.17). Thus, the larger blue, red, and turquoise LCCs resolved into distinct sub-blocks with high modularity and lower global clustering, while the smaller cyan and magenta LCCs behaved as denser cohesive clusters with less sub-clustering.

To characterize the pathway-level function of each of the five LCCs’ full gene sets, we performed over-representation analysis against the Reactome database. The total number of significant Reactome hits at FDR < 0.05 was 58 for red, 142 for blue, 137 for cyan, 14 for turquoise, and 1 for magenta, where the top 5 hits for each module are visualized in Figure 2L (full set of pathways available in Supplementary Table S6). The upregulated blue LCC which was the largest with 1,763 nodes, was most strongly enriched for Neutrophil degranulation, followed by Parasitic Infection Pathways and Leishmania phagocytosis. The upregulated red LCC (660 nodes) was enriched for Hemostasis, Platelet activation, signaling and aggregation, Platelet degranulation, as well as Response to elevated platelet cytosolic Ca² and RHO GTPase Effectors. The downregulated magenta LCC, the smallest with 61 nodes, was solely enriched for immunoregulatory interactions between lymphoid and non-lymphoid cells. The downregulated turquoise LCC (1,598 nodes) exhibited higher FDRs and lower gene ratios than the other modules, indicating a slightly less coherent signal. Top significant pathways were tRNA processing, Class C/3 (metabotropic glutamate/pheromone receptors), defects in vitamin and cofactor metabolism, sensory perception of sweet, bitter, and umami (glutamate) taste, and snRNP assembly. The upregulated cyan LCC (85 nodes) represented an interferon/antiviral network, with Interferon Signaling, Antiviral mechanism by IFN-stimulated genes, and ISG15 antiviral mechanism most significant.

To build a more intuitive understanding of each module’s topology, we visualized the LCC subnetworks and labeled the top hubs per network that ranked highest in composite centrality and were significant in either contrast (CD-vs-DGBI or within-CD association with PCDAI). To resolve each module’s LCC into community structure, we fit an NSBM, which groups nodes into blocks by inferring latent connectivity patterns (nodes assigned to the same block have statistically similar wiring to the rest of the network).

In the blue LCC, 328/1,763 nodes (18.6%) were differentially expressed in at least one model (Figure 3A). Blue nodes were DEGs exclusive to the CD-vs-DGBI model (|log_2_FC| ≥ 0.60, FDR < 0.05), gold nodes were DEGs exclusive to the within-CD model (|log_2_FC| ≥ 0.02, FDR < 0.05), red nodes represented DEGs that were shared across the two models, while gray nodes were not differentially expressed at respective thresholds. The blue module was enriched for diagnostic DEGs, with 164 exclusive to the CD-vs-DGBI model and 149 shared across both models, while only 15 DEGs were exclusive to the within-CD model. These DEGs were not evenly distributed but were significantly concentrated within specific sub-blocks of the network (Supplementary Table S7, z = 13.1, P < 0.001), evident as the dense aggregation of red and blue nodes and edges in the top subclusters of the circular layout. The largest and most dense of these DEG clusters were localized at the top left of the layout, and contained the differentially expressed (DE) hubs *MMP9*, *S100A12*, *FCGR2A*, *FCGR1A*, *TLR4*, *ITGAM*, and *MAPK14*. This cluster connected directly to a second major cluster at the top right of the layout with the most significant hubs being *LTF*, *MPO*, *ARG1*, *TLR8*, and *CASP1*. Smaller DEG blocks were distributed around the layout, represented by *JAK2*, *LRRK2*, *UBE2D1*, *TXN*, *PPARG*, and *H2BC21*. All subclusters remained connected to the main top-left cluster, and the remaining hubs occupied smaller peripheral clusters. All labeled hubs were ranked among the top 120 hubs of the full network (top ∼7%), and their DEG subclusters were strongly represented by Neutrophil degranulation (Figure 3B).

**Figure 3.**
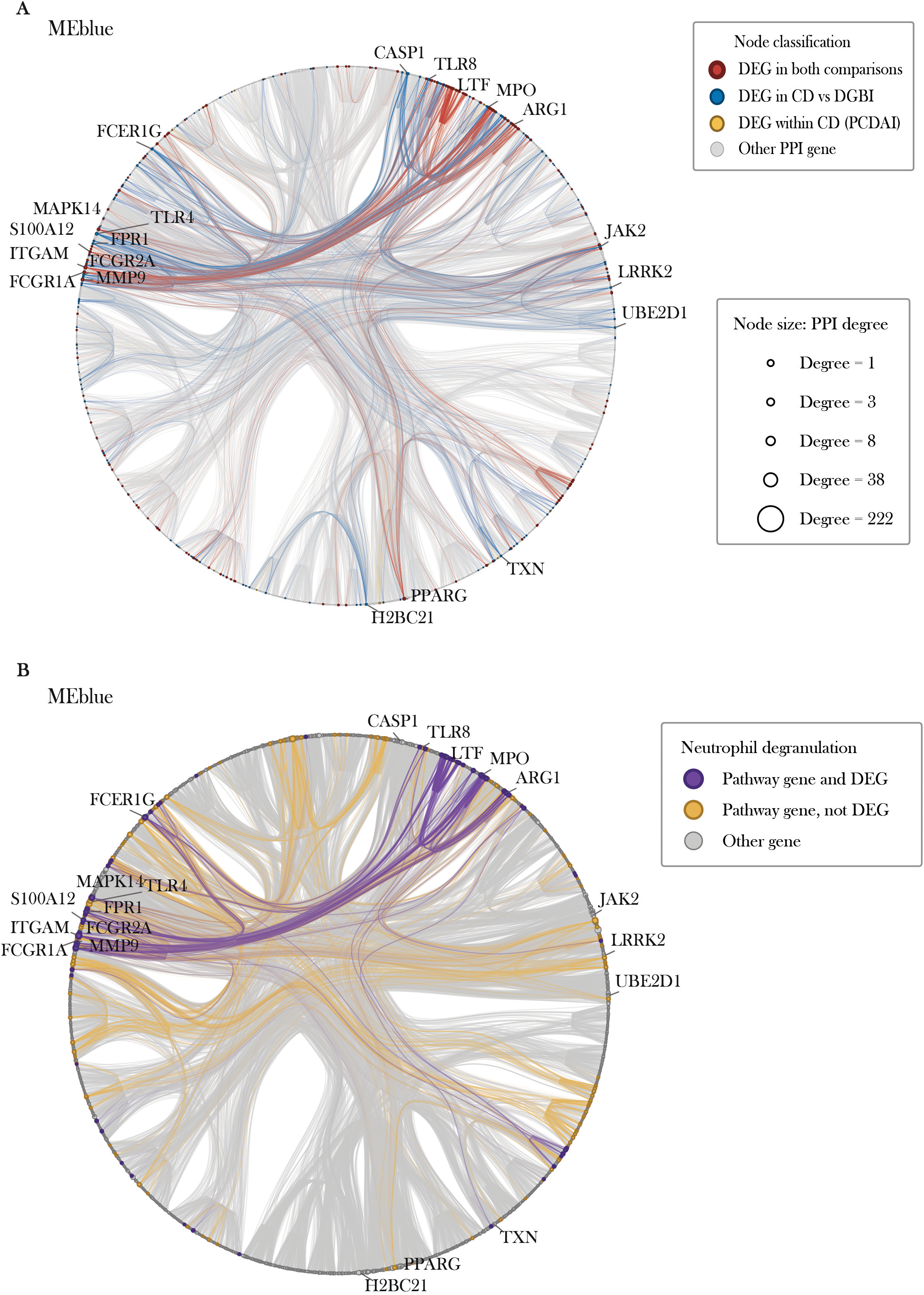
Differentially expressed genes and neutrophil degranulation genes within the blue module protein–protein interaction network. LCC network of the blue module projected from STRING database, shown in the same nested stochastic block model circular layout in both panels. Nodes represent genes, gray lines represent PPI edges filtered by combined score ≥ 500, and node size is proportional to PPI degree (range, 1–222). (A) Nodes colored by differential expression status: genes differentially expressed in both CD-vs-DGBI and within-CD by PCDAI (red), in CD-vs-DGBI only (blue), within-CD by PCDAI only (gold), or not in either DEA model (gray). Edges between differentially expressed genes are highlighted. (B) Genes annotated to the most significant Reactome pathway, Neutrophil degranulation, colored by differential expression status. Pathway genes that were differentially expressed in either CD-vs-DGBI or within-CD model (purple) and pathway genes that were not a DEG (orange). All other genes are shown in gray, and edges involving pathway genes are highlighted. Abbreviations: CD, Crohn’s disease; DEA, differential expression analysis; DGBI, disorders of gut-brain interaction; LCC, largest connected component; PCDAI, Pediatric Crohn’s Disease Activity Index; PPI, protein–protein interaction; STRING, Search Tool for the Retrieval of Interacting Genes/Proteins. **Alt text:** Two network diagrams labeled A and B, each showing the same circular layout of the blue module’s largest connected component, with nodes grouped by community assignment by a nested stochastic block model. Node size is proportional to the number of connections. A shows the nodes colored by differential expression status, where red are genes significant in both models, blue for CD versus DGBI genes only, gold for PCDAI only, and gray for genes not differentially expressed in either, with edges highlighted with the above colors. B highlights the neutrophil degranulation pathway, where pathway genes that are also differentially expressed in either model are colored purple, while nodes colored orange are a pathway gene but not differentially expressed, and gray if neither.

Red’s LCC exhibited a ratio of 158 DEGs to 660 nodes (23.9% differentially expressed), with 94 from the CD-vs-DGBI comparison, 54 overlapping DEGs, and only 10 exclusive to the within-CD model. As in blue, the red module was enriched for diagnostic DEGs, and these were significantly concentrated within the network rather than evenly distributed (Supplementary Table S7, z = 15.1, P < 0.001), forming several subclusters in the left half of the layout and another directly opposite on the right (Figure 4A). The middle block of the right half was enriched predominantly for DEGs shared across both DEA models (red), with *SLC4A1*, *ANK1*, *EPB42*, *ALAS2*, *DMTN*, and *PTGS1* as its most connected hubs. This cluster connected to the opposite half of the network which was enriched for CD-vs-DGBI DEGs (blue), mainly represented by *ITGB3*, *ITGA2B*, *SELP*, *GP6*, *PF4*, *THBS1*, *PDGFRA*, *SERPINE1*, and *EGF*. Smaller clusters were represented by *VCL*, *H2BC11*, *H2BC12*, *FOXO3*, and *PLEK2*. The left-half clusters, which encompassed most diagnostic DEGs, corresponded to Hemostasis (Figure 4B, Figure 2L), consistent with the platelet and coagulation identity of the module. The high modularity of blue and red reflects subnetwork-level perturbation, where particular sub-blocks of genes were differentially expressed, while other sub-blocks of genes exhibited no expression changes.

**Figure 4.**
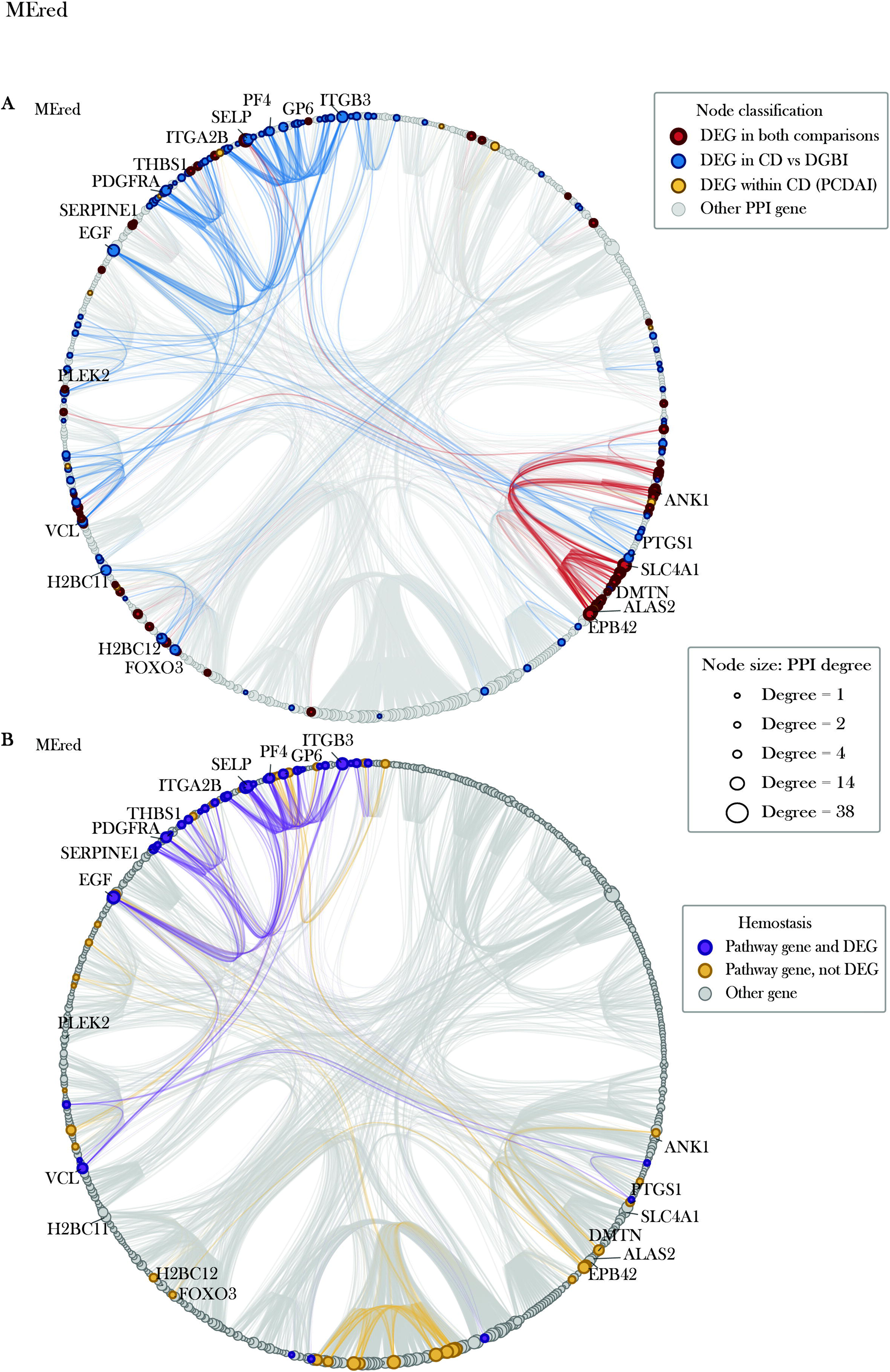
Differentially expressed genes and Hemostasis genes within the red module protein–protein interaction network. LCC network of the red module projected from STRING database, shown in the same nested stochastic block model circular layout in both panels. Nodes represent genes, gray lines represent PPI edges filtered by combined score ≥ 500, and node size is proportional to PPI degree (range, 1–38). (A) Nodes colored by differential expression status: genes differentially expressed in both CD-vs-DGBI and within-CD by PCDAI (red), in CD-vs-DGBI only (blue), within-CD by PCDAI only (gold), or in neither model (gray). Edges between differentially expressed genes are highlighted with corresponding color. (B) Genes annotated to the most significant Reactome pathway, Hemostasis, colored by differential expression status. Pathway genes that were differentially expressed in either CD-vs-DGBI or within-CD model (purple) and pathway genes that were not differentially expressed (orange). All other genes are shown in gray, and edges involving pathway genes are highlighted with respective color. Abbreviations: CD, Crohn’s disease; DEA, differential expression analysis; DGBI, disorders of gut-brain interaction; LCC, largest connected component; PCDAI, Pediatric Crohn’s Disease Activity Index; PPI, protein–protein interaction; STRING, Search Tool for the Retrieval of Interacting Genes/Proteins. **Alt text:** Two network diagrams labeled A and B, each showing the same circular layout of the red module’s largest connected component, with nodes grouped by community assignment by a nested stochastic block model. Node size is proportional to the number of connections. A shows the nodes colored by differential expression status, where red are genes significant in both models, blue for CD versus DGBI genes only, gold for PCDAI only, and gray for genes not differentially expressed in either, with edges highlighted with the above colors. B highlights the hemostasis pathway, where pathway genes that are also differentially expressed in either model are colored purple, while nodes colored orange are a pathway gene but not differentially expressed, and gray if neither.

Turquoise had the lowest DEG proportion at 6% (96 DEGs / 1,598 total nodes), with 49 exclusive to the CD-vs-DGBI model, 20 exclusive to the within-CD model, and 27 shared across both models. Despite the lower DEG proportion, turquoise retained modest but significant DEG block-level concentration (Supplementary Table S7, z = 3.6, P = 0.003). Turquoise’s top DEG hubs were clustered in the blocks at the top of the circular layout: *IFNG*, *TCF7*, *CACNB4*, *FGF9*, *NRCAM*, *WNT7A*, *NOG*, *AXL*, *RAG1*, and *IL23R*, with additional DE hubs distributed around the network including *RBFOX2*, *SARDH*, *FUT2*, *BBS5*, *AK5*, *ENPP1*, *NPR2*, *PDE9A*, and *TLE2* (Supplementary Figure S3A). Notably, the top pathway in the turquoise network was tRNA processing, which contained no DEGs and mapped to a small set of unperturbed blocks (Supplementary Figure S3B). Several other top pathways, including snRNP assembly and metabolism of non-coding RNA, also contained no DEGs, suggesting that housekeeping genes dominated the turquoise module’s top pathways from the LCC-wide enrichment test. To resolve what the DEG-enriched blocks represented, we tested the genes assigned to the most differentially expressed clusters (blocks at the top containing *FGF9*, *CACNB4*, *IFNG*, etc.) of the network for enrichment through GOBP and KEGG, and found they were characterized by T cell receptor signaling (Supplementary Figure S4A) and TGF-beta signaling (Supplementary Figure S4B).

The cyan LCC contained 21 DEGs (24.7% of nodes), of which 19 were exclusive to the CD-vs-DGBI model, 1 exclusive to the within-CD model, and 1 shared across both models (Supplementary Figure S5A). Top DE hubs in the main subcluster at the top half were *EIF2AK2*, *CMPK2*, *EPSTI1*, *SAMD9L*, *GBP6*, *IFI44*, *IFI44L*, *IFIT3*, and *RSAD2*. Two smaller sub-blocks were represented by *GBP1*, *GBP5*, *STAT1*, *PARP9*, *BATF2*, *APOL6*, *CD274*, *PDCD1LG2*, *WARS1*, *ETV7*, and *SERPING1*. Unlike the other 4 modules, DEGs were distributed proportionally across the subclusters (Supplementary Table S7, z = −0.6, P = 0.66). The most enriched pathway, Interferon Signaling, spanned the network but was concentrated in the two smaller subclusters (Supplementary Figure S5B).

Magenta, the smallest network with high clustering and low modularity, had the greatest DEG proportion, with 28 of 61 total nodes differentially expressed (45.9%). In contrast to the other networks, magenta was predominantly a CD severity-associated module, with 23 DEGs exclusive to the within-CD model, 4 shared across both models, and 1 exclusive to the CD-vs-DGBI model (Supplementary Figure S6A). DEGs were also concentrated within the network (Supplementary Table S7, z = 4.3, P = 0.015), although the NSBM only resolved magenta into 2 blocks/clusters where the smaller cluster had no DEGs. The top DE hubs including *GZMK*, *GZMH*, *GZMB*, *IL2RB*, *TBX21*, *KLRD1*, *KIR2DL3*, *KIR2DL1*, *EOMES*, *KLRC1*, *KLRK1*, and *CCL4* were largely characterized by Immunoregulatory interactions between lymphoid and non-lymphoid cells (Supplementary Figure S6B).

### Module-specific classifiers effectively discriminate CD and DGBI samples

We hypothesized that a subset of genes from each co-expression module’s LCC could be used to classify CD from DGBI samples. To test this, we built a 2-stage pipeline (Supplementary Figure S7) composed of (1) identifying the most diagnostic genes in each module; (2) distilling a final panel across each module’s diagnostic subsets. For Stage 1, the candidate classifier genes for each module were 328 DEGs for blue, 158 for red, 96 for turquoise, 28 for magenta, and 21 for cyan. We employed LASSO to select a subset of each module’s candidates, and then LASSO, RF, and SVM models were trained on those selected genes. We refer to these as the locked module panels.

Locked panels were substantially reduced by LASSO, comprising 9 of 328 candidate DEGs for blue, 9 of 158 for red, 14 of 96 for turquoise, 15 of 28 for magenta, and 5 of 21 for cyan, for a total of 52 DEGs across the modules retained (Supplementary Table S8). Positive coefficients indicate higher odds of CD, and negative coefficients indicate lower odds. All 9 blue coefficients were positive, led by *PSTPIP2* (0.479). All 14 turquoise coefficients were negative, led by *CACNB4* (−0.669). Red had 8 positive coefficients and 1 negative coefficient (*RNF152*, - 0.325) and was led by *VCL* (0.475). Magenta was mixed, with 9 positive and 6 negative coefficients, and was led by *GZMK* (−0.869). Among the 5 cyan genes, *CD274* had the largest coefficient (1.072), and *LAMP3* (−0.736) was the only negative coefficient.

All 5 modules discriminated CD from DGBI (Supplementary Table S8). Turquoise performed best, with CV AUCs of up to 0.915 and test AUCs up to 0.955 across the 3 classifiers. Blue followed, with CV AUC up to 0.895 and test AUCs up to 0.914. Red reached CV AUCs of 0.886 and test AUCs of 0.897. Cyan had the lowest CV AUCs (up to 0.852), but its test AUCs reached 0.930 and were exceeded only by those of turquoise. Most magenta candidates were differentially expressed only in the within-CD severity comparison (23 of 28) and had CV AUCs up to 0.891. Its test AUCs were the lowest of the 5 modules and varied the most across classifiers, from 0.803 for RF to 0.869 for LASSO.

Stable-core genes, defined as those selected in at least 900 of 1,000 Stage 1 bootstrap resamples, were few relative to the number of candidates (Supplementary Table S8). Blue and turquoise each contributed a single gene, *PSTPIP2* (stability = 0.902) from 328 blue candidates and *CACNB4* (stability = 1) from 96 turquoise candidates. Cyan also contributed a single gene, *CD274*, with stability = 1. There were 4 genes contributed by both red (*RNF152*, *CHPT1*, *RAP1GAP*, and *CLIC4*) and magenta (*GZMK*, *KLRK1*, *MATK*, and *KLRC1*). All 11 stable-core genes formed the candidate pool for Stage 2. We found network centrality did not reliably predict reproducibility across bootstrapped models. *KLRD1*, the top-ranked hub by composite centrality in magenta, was selected in 631 of 1,000 resamples. In contrast, *MATK*, which was ranked 53rd out of 61 nodes in terms of composite centrality, was selected in 985 resamples. All 4 red stable-core genes ranked 415th or lower out of 660 nodes, and *PSTPIP2*, the only blue stable-core gene, ranked 949th out of 1,763.

### Combined cross-module panel construction and evaluation

To favor sparsity for the final cross-module panel, we recalculated stability frequencies of the 11 module stable genes across another 1,000 bootstraps. Stabilities now ranged from 0.058 to 0.978 (Supplementary Table S9). Four genes exceeded the Stage 2 threshold of 0.85 and formed the final panel. These were *PSTPIP2* from blue (stability = 0.978), *CACNB4* from turquoise (stability = 0.934), *GZMK* from magenta (stability = 0.896), and *CLIC4* from red (stability = 0.871), so 4 of the 5 modules were represented. The next-ranked candidates were both from the red module, *CHPT1* (0.776) and *RNF152* (0.744). No cyan gene was retained, because *CD274*, the only cyan stable-core gene, had a pooled stability of just 0.317 during stage 2 bootstrapping. The remaining magenta stable genes were also infrequently selected across bootstraps. This included *KLRK1*, which had been selected in 993 of 1,000 resamples at the module level (stage 1), but only 58 times in the cross-module (stage 2) bootstraps.

In the final-panel LASSO model, fitted with λ = 0.0565 and an intercept of −1.276, higher expression of *PSTPIP2* (coefficient 0.637) and *CLIC4* (0.415) increased the odds of CD, whereas higher expression of *CACNB4* (−0.423) and *GZMK* (−0.308) decreased them (Supplementary Table S10).

In CV on the training partition of 297 samples, SVM achieved an AUC-ROC of 0.918, LASSO 0.909, and RF 0.901 as shown in Figure 5A and Table 2. AUC-PR ranged from 0.815 to 0.849, sensitivity from 0.769 to 0.795, specificity from 0.922 to 0.941, PPV from 0.785 to 0.822, and NPV from 0.920 to 0.927. On the held-out test partition of 128 samples, AUC-ROC was 0.962 for SVM, 0.961 for LASSO, and 0.927 for RF (Figure 5B). AUC-PR ranged from 0.900 to 0.926, sensitivity from 0.735 to 0.794, specificity from 0.957 to 0.968, PPV from 0.871 to 0.897, and NPV from 0.910 to 0.928.

**Figure 5.**
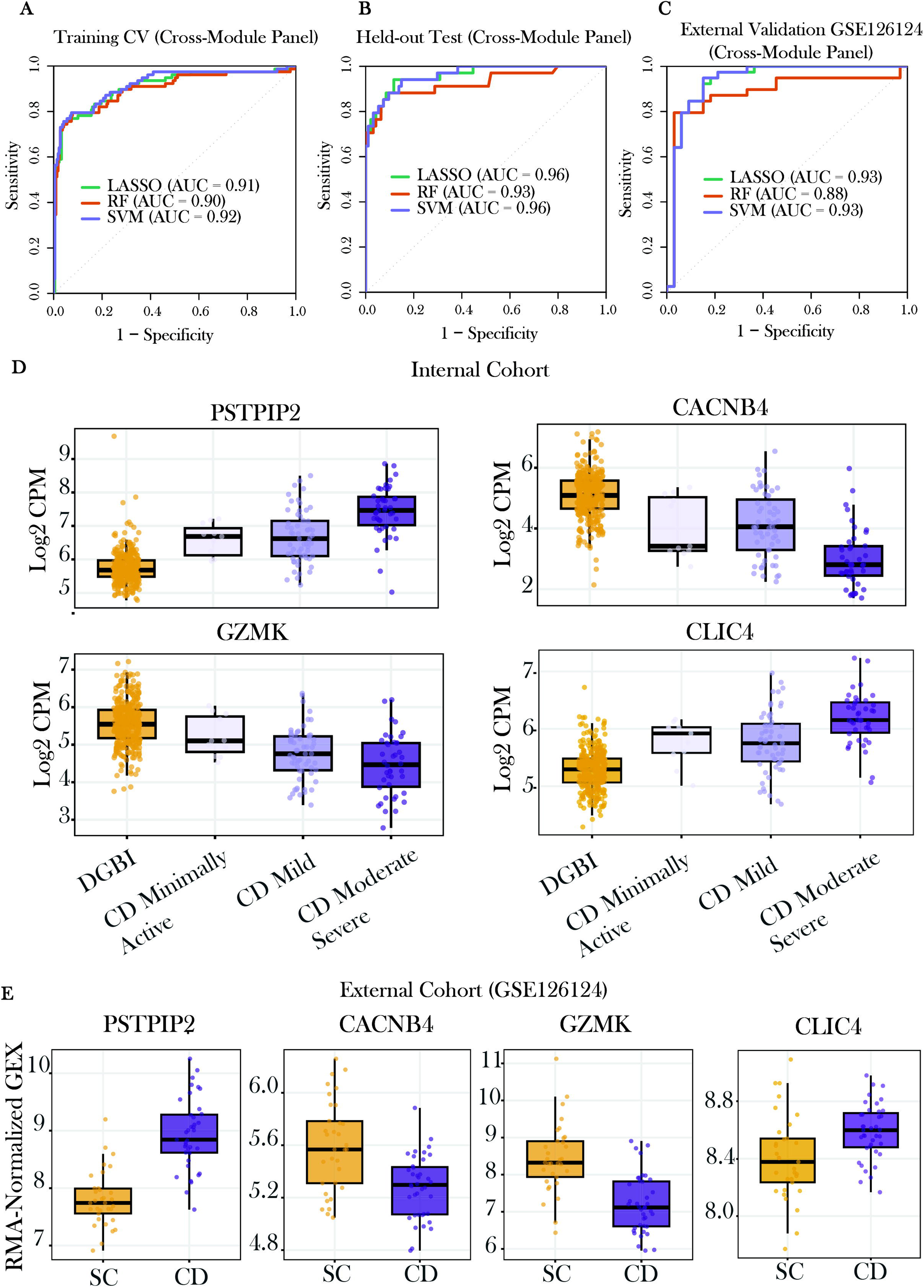
Diagnostic performance and expression of the four-gene whole-blood panel. (A–C) ROC curves for the four-gene panel (*PSTPIP2*, *CACNB4*, *GZMK*, *CLIC4*) classifying CD versus DGBI, using LASSO, random forest (RF), and support vector machine (SVM) classifiers. AUCs are shown in each legend, and the diagonal denotes chance. (A) Out-of-fold predictions from repeated (10 × 10) stratified cross-validation on the internal training partition, averaged per sample across the 10 repeats. (B) A single evaluation on the internal held-out test partition (30%), which was not used for model fitting or stability selection, although candidate genes were derived from the full cohort. (C) External validation in the whole-blood subset of GSE126124 (CD versus SCs; Affymetrix microarray). Models were refit on the full internal cohort, and external data were harmonized by within-cohort gene-wise z-scoring and scored once. (D) Expression of the panel genes in the internal cohort (log_2_ TMM-normalized CPM) across DGBI and CD activity groups defined by PCDAI (minimally-active, mild, moderate-severe). (E) Expression of the panel genes in GSE126124 (RMA-normalized) in SCs and CD. Abbreviations: AUC, area under the curve; CD, Crohn’s disease; CPM, counts per million; DGBI, disorders of gut-brain interaction; LASSO, least absolute shrinkage and selection operator; PCDAI, Pediatric Crohn’s Disease Activity Index; RMA, robust multi-array average; ROC, receiver operating characteristic; SC, symptomatic control; TMM, trimmed mean of M-values. **Alt text:** Graphs labeled A to E. A, B, and C are ROC curves plotting sensitivity versus 1 – specificity for LASSO, random forest, and support vector machine models, for internal cross-validation, held-out test set, and external validation, respectively. Each legend contains AUC values for each model, and a diagonal reference line indicating chance-level performance. D represents the boxplots of log_2_ normalized expression for the four panel genes, PSTPIP2, CLIC4, CACNB4, and GZMK, stratified across DGBI and CD activity groups. E contains boxplots of the same 4 genes, comparing RMA-normalized symptomatic controls and CD samples in the external cohort.

**Table 2.** Classification performance of the 4-gene panel in internal cross-validation, the internal held-out test set and the external validation cohort (GSE126124).

| **Model** | **AUC-ROC (95% CI)** | **AUC-PR** | **Sensitivity** | **Specificity** | **PPV** | **NPV** | **F1 score** | **Balanced accuracy** | **Brier score** |
|---|---|---|---|---|---|---|---|---|---|
| **Internal cross-validation (training partition, n = 297)** |  |  |  |  |  |  |  |  |  |
| LASSO | 0.909 (0.865–0.953) | 0.815 | 0.769 | 0.941 | 0.822 | 0.920 | 0.795 | 0.855 | 0.096 |
| RF | 0.901 (0.854–0.949) | 0.849 | 0.795 | 0.922 | 0.785 | 0.927 | 0.790 | 0.859 | 0.086 |
| SVM | 0.918 (0.877–0.959) | 0.849 | 0.795 | 0.927 | 0.795 | 0.927 | 0.795 | 0.861 | 0.080 |
| **Internal held-out test (n = 128)** |  |  |  |  |  |  |  |  |  |
| LASSO | 0.961 (0.927–0.996) | 0.926 | 0.765 | 0.968 | 0.897 | 0.919 | 0.825 | 0.866 | 0.081 |
| RF | 0.927 (0.865–0.990) | 0.900 | 0.735 | 0.968 | 0.893 | 0.910 | 0.806 | 0.852 | 0.072 |
| SVM | 0.962 (0.930–0.994) | 0.926 | 0.794 | 0.957 | 0.871 | 0.928 | 0.831 | 0.876 | 0.063 |
| **External validation, GSE126124 (n = 76)** |  |  |  |  |  |  |  |  |  |
| LASSO | 0.931 (0.867–0.994) | 0.883 | 0.718 | 0.919 | 0.903 | 0.756 | 0.800 | 0.818 | 0.191 |
| RF | 0.879 (0.790–0.967) | 0.860 | 0.718 | 0.946 | 0.933 | 0.761 | 0.812 | 0.832 | 0.171 |
| SVM | 0.932 (0.869–0.995) | 0.884 | 0.718 | 0.919 | 0.903 | 0.756 | 0.800 | 0.818 | 0.143 |

Thresholds were Youden-optimal cutoffs from internal out-of-fold predictions and were applied unchanged to the held-out and external sets.
Abbreviations: AUC-PR, area under the precision–recall curve; AUC-ROC, area under the receiver operating characteristic curve; CI, confidence interval; LASSO, least absolute shrinkage and selection operator; NPV, negative predictive value; PPV, positive predictive value; RF, random forest; SVM, support vector machine.

All 4 panel genes were present on the external Affymetrix array from GSE126124. SVM and LASSO achieved the highest external AUC-ROCs, 0.932 and 0.931, respectively, while RF reached 0.879 (Figure 5C). AUC-PR ranged from 0.860 to 0.884. At the internally derived thresholds, specificity ranged from 0.919 to 0.946 and PPV from 0.903 to 0.933. Sensitivity was 0.718 for all 3 models, and NPV ranged from 0.756 to 0.761. RF had the highest external balanced accuracy, 0.832 (Table 2).

In our cohort, *PSTPIP2* and *CLIC4* expression were higher in CD compared to DGBI, whereas *CACNB4* and *GZMK* expression were lower (Figure 5D). More specifically, the PCDAI groups revealed that expression within CD varied based on disease activity, particularly within the *GZMK* gene, which consistently decreased across the minimally-active to moderate-severe strata. *CLIC4*, *CACNB4*, and *PSTPIP2* showed little difference across minimally-active and mild groups, with more separation evident in the moderate-severe subgroup. In the external cohort, all 4 genes differed between CD and SCs in the same direction as in the internal cohort (Figure 5E).

### Single gene predictors, leave-one-out analysis, and misclassification rates

*PSTPIP2* had the highest single predictor performance with a univariate CV AUC of 0.891. The other 3 genes performed less well alone (0.806 to 0.838), but still strongly. Removing any single gene from the 4-gene panel lowered the AUC-ROC in every model and partition. During CV, the reduction was no greater than 0.029. On the test partition, it was no greater than 0.046, except for *GZMK* in the SVM model, whose removal reduced the test AUC-ROC by 0.139, to 0.823. Additionally, the PPV was more sensitive to gene removal. It decreased by up to 0.114 in CV after removal of *PSTPIP2* from SVM, and by up to 0.210 on the test partition after removal of *CACNB4* from RF (Supplementary Table S11). By majority vote across the 3 classifiers, CV misclassification was concentrated in samples with mild CD (15 of 41, 36.6%), compared with 12.5% in minimally-active CD, 3.4% in moderate-severe CD, and 6.8% in DGBI. The held-out test partition showed a similar pattern, with minimally-active and mild CD exhibiting the highest misclassification rates, with 66.7% (2 of 3) and 22.2% (4 of 18), respectively (Supplementary Table S12).

## DISCUSSION

In this prospective two-center cohort of 425 treatment-naïve children and young adults, whole-blood transcriptomes separated newly diagnosed CD samples from Rome IV-defined DGBI samples. The program that separated them intensified with disease severity, where all 377 genes shared by the diagnostic and PCDAI models changed in the same direction, and minimally-active CD was most transcriptionally similar to DGBI. A 4-gene panel drawn from 4 co-expression networks (*PSTPIP2*, *CLIC4*, *CACNB4*, and *GZMK*) discriminated CD from DGBI with an AUC of up to 0.96 in held-out samples, and up to 0.93 in an independent pediatric cohort composed of symptomatic controls and CD samples.

Prior whole-blood studies have largely compared IBD with healthy or non-gastrointestinal controls, included treated patients, or were small. In the largest of these, which included children and adults with established IBD (most of whom had received prior therapy), discrimination was high for active pediatric IBD but weak for inactive disease (24). The most comparable study is that of Palmer et al, who prospectively sampled children at diagnostic colonoscopy, including 39 with CD and 39 non-IBD SCs with infectious colitis, juvenile polyps, or functional disorders (26). Their 6-gene blood model reached 80% accuracy (PPV 0.84, NPV 0.72) in an external set of peripheral blood mononuclear cells from healthy adults and adults with treated IBD. Our cohort is more than five times larger and restricts the comparison to CD versus Rome IV-defined DGBI. We used the Palmer et al cohort as our external test set due to its similar pediatric population and usage of whole-blood gene expression, after excluding 2 controls who were later diagnosed with CD. Our panel reached an AUC of up to 0.93, a PPV of up to 0.93, NPV of up to 0.76, and balanced accuracy of up to 0.83. Differences in platform, test population, and prevalence preclude a direct comparison, and we did not evaluate their model in our data. The two panels share no genes, yet all 6 of their panel genes fell within our CD-associated modules. *S100A8*, *SLC2A3*, and *TXNIP* were in the neutrophil (blue) module, where all 3 were indirectly connected to *PSTPIP2* via *IL1RN*. *IL7R* was assigned to the housekeeping and T-cell receptor (turquoise) module and was indirectly connected with *CACNB4* by 3 edges. *UBB* and *ALAS2* were grouped in the hemostasis/platelet (red) module, but each gene was 3 edges away from *CLIC4* and shared no direct neighbors. Taken together, both Palmer’s and our diagnostic panels appear to represent the same neutrophil, lymphoid, and hemostatic pathways indexed through different genes.

The concordance between the diagnostic and severity signatures suggests that CD at presentation differs from DGBI mainly in the magnitude of a systemic inflammatory response. CD samples had higher estimated neutrophil and monocyte fractions and lower lymphocyte fractions. The largest up-regulated network (blue) was significantly associated with neutrophil degranulation, the most reproducible blood pathway across IBD studies (15, 26). The red module added a platelet-activation signal, consistent with the hypercoagulability that peaks around diagnosis in active pediatric IBD (35). The magenta module behaved differently than the other modules, as it was the only one dominated by within-CD DEGs. Specifically, 23 of its 28 DEGs (mainly GZM- and KLR-family genes) declined with increasing PCDAI and were not differentially expressed between CD and DGBI samples. This suggests that loss of circulating effector lymphocytes tracks with severity once CD is established. *GZMK* was an exception, as it was the only magenta gene differentially expressed between CD and DGBI without being associated with the PCDAI, yet was still retained in the final panel. Within the blue, red, turquoise, and magenta networks, stochastic block modeling showed that DEGs were concentrated in discrete, interconnected sub-blocks rather than dispersed across the network. This suggests that CD perturbs specific subnetworks within each program rather than the program as a whole.

A notable finding was that sampling several pathways matched or improved predictions compared with sampling one pathway in depth. The blue and red modules had coherent functional identities, Neutrophil degranulation and Hemostasis, and the strongest differential expression signals. Yet neither module’s own locked panel (9 genes each, with test AUC up to 0.91 and 0.90) matched the 4-gene cross-module panel with test AUC of up to 0.96. Moreover, the genes within a single program (ie, Neutrophil degranulation) were highly collinear, which was reflected by LASSO rotating among blue-module genes across bootstraps. Additionally, the *PSTPIP2* model alone reached CV AUC of 0.89, while the full blue module only increased this by less than 0.01 CV AUC with 8 more genes. This pattern suggested that adding more genes does not necessarily add more information. Proxies for separate programs, by contrast, contribute complementary signal. The same logic may explain why turquoise, whose network lacked one dominant biological process, produced the best single-module panel (test AUC up to 0.95) which the 4-gene cross-module panel matched with less than a third as many genes. The final panel reflects this breadth. *PSTPIP2* is a known myeloid adaptor that restrains neutrophil IL-1β and NADPH oxidase activity (36). The hemostasis (red) module contributed *CLIC4*, which is a lipopolysaccharide-inducible intracellular chloride channel that promotes macrophage inflammatory cytokine production (37). *GZMK* and *CACNB4* both index the downregulated lymphocyte program, where GZMK+ CD8+ T cells were shown to accumulate in inflamed CD mucosa (38). Removing any single gene reduced AUC by no more than 0.03 during CV and 0.05 during testing, except for *GZMK* in the SVM model (0.14 drop in AUC). Our panel should therefore be interpreted as a set of pathway proxies rather than as candidate mechanisms.

The panel’s operating characteristics favor a rule-in role due to its high specificity. In the external cohort, specificity reached 0.95 but sensitivity did not exceed 0.72, and misclassified CD samples were concentrated in mild and minimally-active disease. A positive result could therefore justify expedited endoscopy, whereas a negative result could not exclude mild CD. Thresholds derived on RNA-seq will need recalibration on a targeted assay, and the panel’s value beyond fecal calprotectin and CRP remains to be established. This study has limitations as well. Candidate genes were derived from all 425 samples from WGCNA and before the training/test split, so held-out test estimates are likely optimistic. The external cohort, which played no role in gene selection, provides the least biased estimate. The STRING database restricted our network analysis to protein-coding genes, so noncoding and poorly annotated genes were lost from every module, including those that were differentially expressed. PCDAI is weighted toward symptoms that partly overlap with DGBI, and endoscopic activity was not modeled. Additionally, only 11 patients had minimally-active CD, reducing power to detect differences in the mildest of CD cases. CRP was not collected systematically and was missing in 102 participants, so we did not benchmark our panel against it. Because participants with ulcerative colitis and celiac disease were excluded during study recruitment, the panel’s performance against other inflammatory conditions, other than the small number of infectious colitis SCs in the external cohort, remains untested. Future work should test the panel against fecal calprotectin and CRP in a referral population that includes other inflammatory diagnoses, using a targeted qPCR assay for *PSTPIP2*, *CLIC4*, *CACNB4*, and *GZMK*. If its performance is confirmed, this panel would be suitable for a low-cost, point-of-care blood test. The findings presented in this study should be followed up using paired biopsies with endoscopic scoring, single-cell profiling, and longitudinal sampling to determine the panel’s prognostic value.

At presentation, children and young adults with CD carry a whole-blood signature of neutrophil and platelet activation and lymphocyte depletion that distinguishes them from peers with DGBI and scales with severity. A 4-gene panel sampling these programs performed well in an independent cohort and may help prioritize symptomatic children for further evaluation.

## Data Availability

All data produced in the present study are available upon reasonable request to the authors.

https://www.ncbi.nlm.nih.gov/geo/query/acc.cgi?acc=GSE126124

## ACKNOWLEDGEMENTS

We thank the participants and their families, and the clinical research teams at NYU Langone Health and the Children’s Hospital of Philadelphia. We also want to thank the Leona M. and Harry B. Helmsley Charitable Trust whose donations made this research possible. This research was supported in part by the NYU High-Performance Computing Facility at NYU Langone Health.

## AUTHORS’ CONTRIBUTIONS

C.L., K.M., D.F., J.L., and L.T. conceived the study. C.L. performed all bioinformatic analyses, generated the figures and tables, and drafted the manuscript. K.M. and J.L. contributed to writing the manuscript. P.K.M., V.K., Y.A., and J.O.-M. collected and curated the covariate data. P.K.M., L.A., D.P., and J.O.-M. managed the project. P.Z. and P.M. prepared the samples and performed RNA sequencing. J.L. and L.T. obtained funding and supervised the study. All authors critically reviewed and edited the manuscript and approved the final version.

## DATA AVAILABILITY STATEMENT

The external validation data are publicly available from the Gene Expression Omnibus under accession GSE126124 (26). The whole-blood RNA-seq data generated in this study are available from the corresponding author on reasonable request.

## Abbreviations

AUC: area under the curve
AUC-PR: area under the precision–recall curve
AUC-ROC: area under the receiver operating characteristic curve
BMI: body mass index
CD: Crohn’s disease
CHOP: Children’s Hospital of Philadelphia
CPM: counts per million
CRP: C-reactive protein
CV: cross-validation
DCM: differentially co-expressed module
DE: differentially expressed
DEA: differential expression analysis
DEG: differentially expressed gene
DGBI: disorders of gut-brain interaction
FDR: false discovery rate
GI: gastrointestinal
GOBP: Gene Ontology Biological Process
IBD: inflammatory bowel disease
IBS: irritable bowel syndrome
KEGG: Kyoto Encyclopedia of Genes and Genomes
LASSO: least absolute shrinkage and selection operator
LCC: largest connected component
log_2_FC: log_2_ fold change
ME: module eigengene
NPV: negative predictive value
NSBM: nested stochastic block model
NYU: NYU Langone Health
PCA: principal component analysis
PCDAI: Pediatric Crohn’s Disease Activity Index
PPI: protein–protein interaction
PPV: positive predictive value
RF: random forest
RNA-seq: RNA sequencing
SC: symptomatic control
STRING: Search Tool for the Retrieval of Interacting Genes/Proteins
SVM: support vector machine
TMM: trimmed mean of M-values
WGCNA: weighted gene co-expression network analysis.

