## Supplementary Figures S1-S7, Supplementary Methods for "A Whole-Blood Four-Gene Panel Distinguishes Pediatric Crohn’s Disease from Disorders of Gut-Brain Interaction"

***Study Design***

This study drew on participants from a prospective observational cohort. Between September 2019 and June 2024, individuals aged 5–25 years presenting with undiagnosed gastrointestinal symptoms at NYU Langone Health, New York, NY (NYU) or the Children’s Hospital of Philadelphia, Philadelphia, PA, (CHOP) were enrolled. Patients were ultimately diagnosed with (a) CD by standard radiologic and histologic criteria or (b) DGBI according to the Rome IV criteria (1). Individuals with biopsy-confirmed ulcerative colitis, celiac disease, or CD who received treatment prior to sample collection were excluded. All CD diagnoses were independently reviewed by the site study team and confirmed using established criteria that incorporated clinical history, physical examination, laboratory testing, and endoscopic and/or radiologic evaluation. Disease activity in CD was assessed with PCDAI, and patients were stratified into minimally-active (PCDAI ≤ 10), mild (10 < PCDAI ≤ 30), or moderate–severe (30 < PCDAI) categories based on accepted thresholds from previous studies (2, 3). These groups were used to summarize the CD population but not utilized for downstream modeling.

Whereas the parent protocol included collection of multiple biospecimens, the present analysis focused exclusively on total RNA from whole blood. Across the study period, 438 participant samples were sequenced. Thirteen samples were excluded per prespecified QC/eligibility criteria, yielding a final analytic cohort of 425 samples (112 CD, 313 DGBI) used for all downstream analyses. All participants (and guardians, where appropriate) provided written informed consent, and study procedures were approved by the Institutional Review Boards of the NYU Grossman School of Medicine and the Children’s Hospital of Philadelphia.

***Whole-Blood RNA-seq***

Total RNA was extracted from whole blood and quality was verified by Bioanalyzer trace. Library preparation was performed using the Tecan RNA library preparation kit, which is compatible with a wide range of RNA integrity. The protocol included ribosomal RNA (rRNA) depletion and integrated DNase treatment. For each sample, 5 ng of total RNA was used as input, and 10 PCR cycles were applied during library amplification. Library concentrations were quantified by quantitative PCR (qPCR), and samples were pooled to achieve uniform sequencing depth. Sequencing was performed on the Illumina NovaSeq X Plus platform, generating 2 × 51 bp paired-end reads, with one plate loaded per flow cell. Fresh reagents were used for each plate run.

***RNA-seq Preprocessing***

In total, 438 bulk RNA patient samples were processed on NYU Langone’s high-performance computing (HPC) cluster. Each batch was processed using nf-core/rnaseq v3.17.0 (10.5281/zenodo.1400710) (4) under Nextflow v24.10.3 with a custom HPC configuration. Default pipeline parameters were used unless otherwise specified. Briefly, reads were merged across lanes with cat v9.5, quality-checked using FastQC v0.12.1 before and after trimming with Trim Galore v0.6.10, and rRNA sequences were identified against the default database and removed using SortMeRNA. STAR v2.7.11b aligned reads to GRCh38 using the most current Ensembl GTF/FASTA (v113). Transcript quantification was performed with Salmon v1.10.3. MultiQC v1.25.1 summarized quality-control metrics.

Transcript-level quantifications were imported into R v4.4.1 (5) using tximport v1.32.0 and aggregated to the gene level. Thirteen samples with STAR alignment < 85% were removed. Counts were processed with edgeR v4.2.2 across CD and DGBI samples. The hemoglobin genes HBAP1, HBA1, HBA2, HBB, HBBP1, HBD, HBE1, HBG1, HBG2, HBM, HBQ1, HBZ, HBZP1 and pseudogenes were excluded. Low-signal genes were filtered with filterByExpr based on the minimum library size and CD group size. Library-size scaling factors were computed via Trimmed Mean of M-values (TMM), and counts were normalized accordingly. TMM-normalized counts were used to explore batch effects, where the limma package was utilized to evaluate covariate-associated variance while preserving the biological variable of interest (6).

***Immune Cell Estimation***

Immune cell composition was estimated from the log-transformed TMM-normalized expression matrix using CIBERSORT with the LM22 signature matrix (7). Wilcoxon rank-sum tests were performed to evaluate differences in inferred immune-cell proportions between the CD and DGBI groups.

***Differential Expression Analysis***

Two models were evaluated: (a) a diagnostic comparison of CD vs DGBI samples, and (b) a within-CD model associating gene expression with PCDAI. For both, TMM-normalized counts were transformed using voom with quality weights. Gene expression was modeled as a function of (a) CD diagnosis or (b) PCDAI, adjusting for sequencing batch, sex, secondhand smoke exposure, sample recruitment site, and age for both models. Differentially expressed genes (DEGs) were identified with lmFit and eBayes from the limma package (6). Significance thresholds were model-specific: in the CD-vs-DGBI model, DEGs had |log_2_FC| ≥ 0.60 and false discovery rate (FDR) < 0.05; in the within-CD model, DEGs had |log_2_FC| ≥ 0.02 and FDR < 0.05.

To identify shared transcriptional signals which highlight both diagnosis and disease severity, we compared significant gene sets from the two models using a Venn diagram and carried overlapping DEGs forward to pathway enrichment. Venn diagrams were constructed using ggVennDiagram. Functional over-representation analysis of the overlapping DEGs was performed using clusterProfiler (8) using all 15,444 genes as the background universe and Benjamini-Hochberg adjusted FDR < 0.05. Gene sets were drawn from Kyoto Encyclopedia for Genes and Genomes (KEGG), and Gene Ontology-Biological Process (GOBP). Curated results across databases were visualized with dot plots to provide an integrated overview of transcriptional programs perturbed in CD.

To prepare for downstream visualization, network analysis, and classification, we fit a linear model that included all covariates except CD diagnosis and extracted model residuals. These residuals represented expression values with covariate effects removed, emphasizing CD-associated biological signal. Heatmaps were generated from residual counts using pheatmap with row-scaling, and hierarchical clustering using Euclidean distance and complete linkage was applied to rows and columns.

***Weighted Gene Co-Expression Network Analysis***

Weighted Gene Co-Expression Network Analysis (WGCNA) was employed to construct gene co-expression networks by identifying modules of highly correlated genes across samples (9). This technique facilitated the detection of biologically relevant gene clusters associated with clinical traits, which enabled insights into coordinated gene expression patterns in the CD and DGBI samples. WGCNA v1.73 was performed on residual counts to account for known covariates (age, sex, secondhand smoke exposure, sample recruitment site, and sequencing batch) prior to network construction. The analysis aimed to identify co-expression modules and central gene hubs whose coordinated activity differed between CD and DGBI groups. Soft-thresholding powers from 1–20 were evaluated to approximate the best fit for scale-free topology. Networks were built as signed adjacency and topological overlap (TOM). Modules were detected in a single call to the blockwiseModules function with the following settings: signed network type and signed TOM type; minimum module size = 30 genes; deepSplit = 3; a module-merging cut height = 0.25, corresponding to the merging of modules whose eigengenes were correlated at r >= 0.75; and a reassignment threshold of 1e-6. Branch identification used the dynamic hybrid tree-cut algorithm (dynamicTreeCut), and the partitioning-around-medoids stage was not constrained to the dendrogram (pamRespectsDendro = FALSE). Detected modules were assigned standard color labels (numericLabels = FALSE), with genes not assigned to any module placed in the grey module.

**Differential Module Expression Analysis and overlap with DEGs**

Each module was summarized by its module eigengene (ME), the first principal component of standardized module gene expression, which served as a composite value that captures the dominant expression pattern of the entire gene group (9). MEs were correlated with clinical/technical variables including CD diagnosis and PCDAI group status using Pearson correlation to assess module-trait associations. To prioritize differentially co-expressed modules (DCMs) for downstream network analysis, module genes were compared with DEGs identified in the within-CD disease activity analysis and the CD-vs-DGBI analysis. Differential expression was defined as previously described above, where FDR < 0.05 with an absolute |log_2_FC| >= 0.02 for the within-CD analysis (PCDAI) and |log_2_FC| >= 0.60 for CD-vs-DGBI. For each module, the numbers and proportions of genes differentially expressed in each comparison, either comparison (the union), and both comparisons (the intersection) were summarized. Modules in which more than 5% of genes were differentially expressed in at least one comparison were retained for downstream analyses – these modules were denoted as DCMs.

***STRING Reconstruction of Protein-Protein Interaction (PPI) Network***

Network analysis was conducted separately for each selected DCM. Protein identifiers and associations were obtained using STRINGdb and STRING version 12.0. Analyses were restricted to Homo sapiens (NCBI taxonomy identifier 9606). For each module, a separate STRING protein association network was constructed using a minimum combined association score of 500 on the STRING 0–1000 scale. STRING associations integrate evidence from multiple sources and may represent direct physical interactions or indirect functional associations. Therefore, the resulting graphs were treated as protein association networks rather than exclusively as experimentally validated physical PPI networks.

An undirected graph was constructed for each module, including mapped proteins without retained interactions as isolated nodes, and duplicate and self-interactions were removed. The largest connected component (LCC) was extracted from each graph. Network structure was summarized using the number of nodes and edges, the proportion of mapped nodes contained in the LCC, the global clustering coefficient, and Louvain modularity weighted by STRING combined scores. Edges excluded during LCC extraction were calculated as the difference between the full-network and LCC edge counts.

***Overrepresentation analysis of LCC***

Reactome pathway over-representation analysis was performed separately for the genes in the LCC of each module-specific PPI. The background universe consisted of all 15,444 genes used in this dataset. Pathway enrichment was assessed using a hypergeometric test implemented in ReactomePA (10), and P-values were corrected within each module using the Benjamini–Hochberg false discovery rate procedure. Pathways with FDR-adjusted P-values < 0.05 were considered significant.

***Network Visualization/Modeling***

Downstream network analyses were performed in Python v3.12 using gprofiler-official v1.0.0, matplotlib v3.8.4, networkx v3.3, graph-tool v2.59, numpy v1.26.4, pandas v2.2.2, and requests v2.31.0. To resolve each DCM’s LCC into hierarchical community structure, we fit a nested (hierarchical) stochastic block model (NSBM) to each LCC using graph-tool. Blocks were inferred by minimizing the description length of the degree-corrected NSBM (minimize_nested_blockmodel_dl), an information-theoretic criterion that identifies groups of nodes with statistically similar connectivity while penalizing model complexity, thereby providing a principled, multi-resolution partition without a user-specified block count. The finest (level-0) partition was retained allowing for visualization of biologically meaningful substructures within the network (11).

***Blockwise-DEG Test***

To test whether DEGs were concentrated within specific blocks rather than dispersed across the LCC, we quantified the deviation of the observed per-block DEG distribution from that expected under the module-wide DEG rate using a *χ²*-type statistic,

$$S = \sum_{b\mathcal{\in B}} \frac{\left( k_{b} - e_{b} \right)^{2}}{e_{b}}$$

where the sum runs over *ℬ*, the set of *B* blocks in the finest (level-0) NSBM partition, *k_b_* is the observed number of DEGs in block *b*, and *e_b_* is the corresponding expected count,

$$e_{b} = n_{b} p, p = \frac{k}{n}$$

with *n_b_* the size of block *b*, *p* = *k* / *n* the module-wide DEG proportion, *k* the total number of DEGs, and *n* the total number of nodes in the LCC. Blocks with zero expected DEGs (i.e. when *p* = 0) were excluded from the summation.

Under the null hypothesis, DEG status is independent of block membership, such that DEGs are distributed across blocks in proportion to block size. The null distribution of *S* was generated by randomly permuting DEG labels across all LCC nodes (*n_perm_* = 10,000 permutations), which preserves the block partition, the block sizes, and the total number of DEGs (k) while randomizing which nodes are labeled as DEGs. The per-block counts *K_b_* arise from wherever the labels fall. Blocks were treated as fixed, so the test assesses DEG localization given the inferred partition. A one-sided permutation P-value was computed with a plus-one correction to avoid a value of zero,

$$P = \frac{\sum\left[ S^{*} \geq S_{obs} \right] + 1}{n_{perm} + 1}$$

where *S^*^* denotes the statistic computed on a permuted labeling and *S_obs_* the observed statistic. A standardized effect size was reported as

$$z = \frac{S_{obs} - \mu_{null}}{\sigma_{null}}$$

where *μ_null_* and *σ_null_* are the mean and standard deviation of the null distribution. Modules whose LCC resolved into a single block were not testable.

***Hub Genes***

Hub genes represent critical nodes within the interaction network whose high connectivity places them at the center of module function, and their altered expression can disrupt network integrity and produce substantial shifts in biological function (12). Because such nodes are essential for maintaining network connectivity, they have been proposed as candidate drug targets, sites of disease-associated mutation, and features for disease prediction (13, 14). Four complementary centrality measures were computed for every node on each module’s PPI: degree, normalized betweenness centrality, PageRank (damping factor 0.85), and k-core index. To avoid reliance on any single measure, genes were ranked on each metric and a composite hub rank was derived as the mean of the four ranks, re-ranked so that a rank of one denoted the most consistently central gene across the four metrics. This consensus strategy reduced the influence of measure-specific artifacts, including the tendency of STRING node degree to reflect annotation and literature bias rather than biological centrality. Each gene was labeled with its differential expression statistics from both DEA models: CD-vs-DGBI, and within-CD association with the PCDAI. The adjusted p-values of each model were ranked separately. The highest-ranked hub genes per module, together with their centrality and differential expression statistics, were used to label each LCC with their respective top hubs.

***Classification of CD and DGBI Samples***

The classification pipeline is illustrated in Supplementary Figure S7.

**Cohorts and outcome**

The internal cohort comprised 425 participants with whole-blood RNA-seq after quality control, of whom 313 had DGBI and 112 had CD, a CD prevalence of 0.263. CD was the positive class. The external cohort was the whole-blood subset of GSE126124, profiled by Affymetrix microarray, comprising 39 patients with CD and 37 non-IBD symptomatic controls (SC), a CD prevalence of 0.513. The external comparator group and the expression platform therefore both differed from the internal cohort.

**Candidate features**

WGCNA and differential expression analysis were performed on all 425 internal samples before the training/test split. Five co-expression modules projected onto STRING, filtered to their LCC, were each reduced to their DEGs with the prespecified cutoffs above. Thus, a module gene was carried forward if it was differentially expressed between CD and DGBI with |log_2_FC| >= 0.60 and FDR < 0.05, or if it was associated with severity within CD, with PCDAI modeled as a continuous predictor and |log_2_FC| >= 0.02 and FDR < 0.05.

**Data partitioning**

After candidate-gene definition, samples were split 70/30 into a training partition of 219 DGBI and 78 CD samples and a held-out test partition of 94 DGBI and 34 CD samples. The split was stratified by severity subgroup with fixed seed, and the same stratification was used for outer CV folds and stability-selection bootstraps. Within the modeling pipeline, the held-out test partition was used only for final evaluation and never for gene selection, model fitting, hyperparameter tuning, or threshold derivation.

**Stage 1: per-module nested cross-validation and locked module models**

Each module was analyzed independently on the training partition by repeated nested CV with 10 outer folds and 10 repeats. These module-specific analyses assessed each module’s ability to distinguish CD from DGBI using its own set of DEGs. Genes were standardized using parameters estimated on the outer training fold only, which were then applied to the corresponding outer validation fold. Within each outer training fold, LASSO logistic regression (glmnet, α = 1) was tuned by inner 10-fold CV, with λ selected as lambda.1se of the cross-validated binomial deviance over a 250-value automatically generated sequence using glmnet’s default lambda.min.ratio. Genes with nonzero coefficients were passed to RF and SVM, which were each tuned by inner 5-fold grid search. RF used ranger with 1,000 trees, mtry chosen from 0.10p, 0.25p, 0.50p, and √p, where p is the number of genes, supplemented with the integers 1 to min(5, p), and min.node.size chosen from 1, 5, and 10. The SVM used e1071 with a radial kernel, cost chosen from 0.1, 1, 10, and 100, and γ chosen from “scale,” defined as 1/[p × var(X)], 0.01, 0.1, and 1. Because gene selection was repeated within each fold, the gene set underlying the cross-validated estimates varied across folds. Out-of-fold predicted probabilities were averaged per sample across the 10 repeats, and all CV metrics and confidence intervals were computed on this single averaged prediction per sample. The nested-CV selection frequency of each gene was the proportion of outer fits in which LASSO selected it.

After nested CV, a locked model was fitted once per module on the full training partition. LASSO, tuned as above, defined the module’s locked panel—the genes with nonzero coefficients in the module’s own final fit. RF and SVM were then fitted to the locked-panel genes using the exhaustive grids described above, and all three locked module models were scored once on the held-out test partition.

**Stability selection**

Here, we define a gene’s stability as the fraction of the N bootstrap LASSO fits in which that gene was selected. Stability selection drew 1,000 bootstrap resamples of the training partition, stratified by severity subgroup. In each resample, LASSO with α = 1 was fitted with λ tuned by inner 5-fold CV, and a gene’s selection frequency was the proportion of resamples in which its coefficient was nonzero. At Stage 1, stability selection ran alongside each module’s LASSO. It characterized reproducibility and defined the Stage 2 candidate pool, but did not limit the module’s own model, so a module’s locked panel could contain genes below the stability threshold. To favor sparsity for he cross-module panel in Stage 2, genes with a stability frequency of at least 0.90 formed a module’s stable core - that is, genes selected in at least 900/1,000 bootstraps. In highly collinear modules, LASSO rotates its selection among correlated genes across resamples, so few genes may clear this threshold even when the module carries a strong signal. Thus, the most stably-selected predictors from each module, representing distinct perturbed biological pathways, were subsequently pooled into a sparse cross-module panel designed to improve the predictive ability of using just one module while minimizing the number of required genes.

**Stage 2: combined cross-module panel**

After the stable cores of all module-LCC’s were pooled, stability selection was repeated on the cross-module candidates with the same settings, and genes with a pooled frequency of at least 0.85 were retained. At this stage, stability selection therefore preceded and defined the input to the final model fit, and the reported pooled frequencies are the membership-defining frequencies.

LASSO, RF, and SVM were then fitted to the twice-stability filtered genes on the training partition as locked models. LASSO λ was selected as in the above description in stage 1. RF and SVM were tuned by exhaustive inner 10-fold grid search. For RF, mtry was chosen from 0.05p, 0.10p, 0.15p, 0.20p, 0.25p, 0.30p, 0.40p, 0.50p, 0.60p, 0.70p, 0.80p, 0.90p, and 1.00p, √p, and log_2_ p, supplemented with the integers 1 to min(5, p), and min.node.size from 1, 2, 3, 5, 7, 10, 15, 20, and 30, which gave 36 unique combinations for 4 genes. For SVM, cost was chosen from 0.001, 0.01, 0.1, 0.5, 1, 2, 5, 10, 25, 50, 100, 250, 500, and 1000, and γ from “scale,” 0.001, 0.005, 0.01, 0.05, 0.1, 0.25, 0.5, 1, 2, and 5, giving 154 combinations. The selected hyperparameters were λ = 0.0565 (log λ = −2.873) for LASSO, mtry = 1 and min.node.size = 1 for RF, and cost = 0.001 and γ = 0.005 for SVM.

Because module-level and pooled stability selection were performed on the full training partition prior to the combined-panel cross-validation, the genes eligible for the final panel were not re-selected within each cross-validation fold. The combined-panel CV estimate is therefore not fully nested with respect to feature selection and is optimistically biased. For the held-out test, gene selection, stability selection, model fitting, hyperparameter tuning, standardization, and threshold derivation used the training partition only. However, the candidate DEGs and co-expression modules were defined on all 425 internal samples, including the 128 held-out test samples, so the test data informed the candidate gene pool, although no model was fitted to them. The external cohort played no part in feature definition, model fitting, tuning, or threshold selection. GSE126124 was standardized gene-wise using only its own expression distribution, without outcome labels, and the thresholds applied to it were derived from out-of-fold predictions on the internal cohort.

**Coefficients and feature importance**

Two distinct sets of LASSO coefficients are reported. Module-level coefficients come from each module’s own locked LASSO fit in Stage 1 and describe that module’s locked panel (Supplementary Table S8). Final-panel coefficients come from the locked Stage 2 LASSO fitted to the cross-module-stable genes, and only these describe the reported panel model (Supplementary Table S10). A gene’s module-level and final-panel coefficients therefore differ in magnitude and should not be interchanged. All coefficients are on the standardized expression scale, and a positive sign indicates higher odds of CD.

**Performance metrics and thresholds**

AUC-ROC with 95% confidence intervals was computed with pROC (DeLong method), and AUC-PR with PRROC. Sensitivity, specificity, PPV, NPV, F1 score, balanced accuracy, and the Brier score, defined as the mean squared error of predicted probabilities, were also reported. All operating thresholds were Youden-optimal and were derived from out-of-fold cross-validated predictions on internal data only. None was derived from resubstituted, in-sample predictions or from the cohort being evaluated. Two distinct thresholds were derived for 2 evaluation sets. For the held-out test partition, the threshold was estimated from the out-of-fold CV predictions on the training partition, to which the held-out test data did not contribute; the resulting thresholds were 0.293 for LASSO, 0.345 for RF, and 0.350 for SVM. For external validation, the scored model was the refit on all 425 internal samples, a model distinct from the training-partition model (same gene panel, different coefficients and thresholds). A new threshold was therefore re-estimated from out-of-fold CV predictions on all 425 internal samples, with the gene set fixed and models refit within each fold. These thresholds were 0.287 for LASSO, 0.342 for RF, and 0.463 for SVM, and external outcomes were never used to select them. Threshold-free metrics, AUC-ROC and AUC-PR, do not depend on either threshold.

**Single-gene and leave-one-out analyses**

Each panel gene was evaluated alone by univariate logistic regression under the same out-of-fold CV scheme on the training partition, so that single-gene AUCs are directly comparable with the panel’s CV AUCs. For the leave-one-out analysis, each gene was removed in turn and the full analysis, covering internal CV and the held-out test, was repeated on the remaining 3 genes. The change in each metric relative to the full panel is reported (Supplementary Table S11).

**External validation of GSE126124**

Samples were subset from GSE126124, specifically filtering to the 39 CD and 37 symptomatic control samples. Samples were profiled on the Affymetrix Human Gene 1.0 ST array (GPL6244) and RMA-normalized. Controls had juvenile polyps, infectious colitis, or functional intestinal disorders, and 2 controls later diagnosed with CD were excluded. The final gene set was held fixed, and the LASSO, RF, and SVM models were refit on the full internal cohort of 425 samples. Coefficients, hyperparameters, and the operating threshold were all derived from internal data. Nothing was fitted, tuned, or selected on the external cohort, which was accessed once for scoring. Because absolute expression scales are not comparable between RNA-seq and microarray, each gene was z-scored within each cohort separately. All of our panel genes were present on the array.

**Misclassification by severity subgroup**

Samples were classified by majority vote across the 3 models at their internally derived out-of-fold Youden thresholds. Misclassification rates were tabulated by severity subgroup for out-of-fold CV predictions and for held-out test predictions (Supplementary Table S12). Subgroup labels were never used as model inputs.

**SUPPLEMENTARY FIGURES**

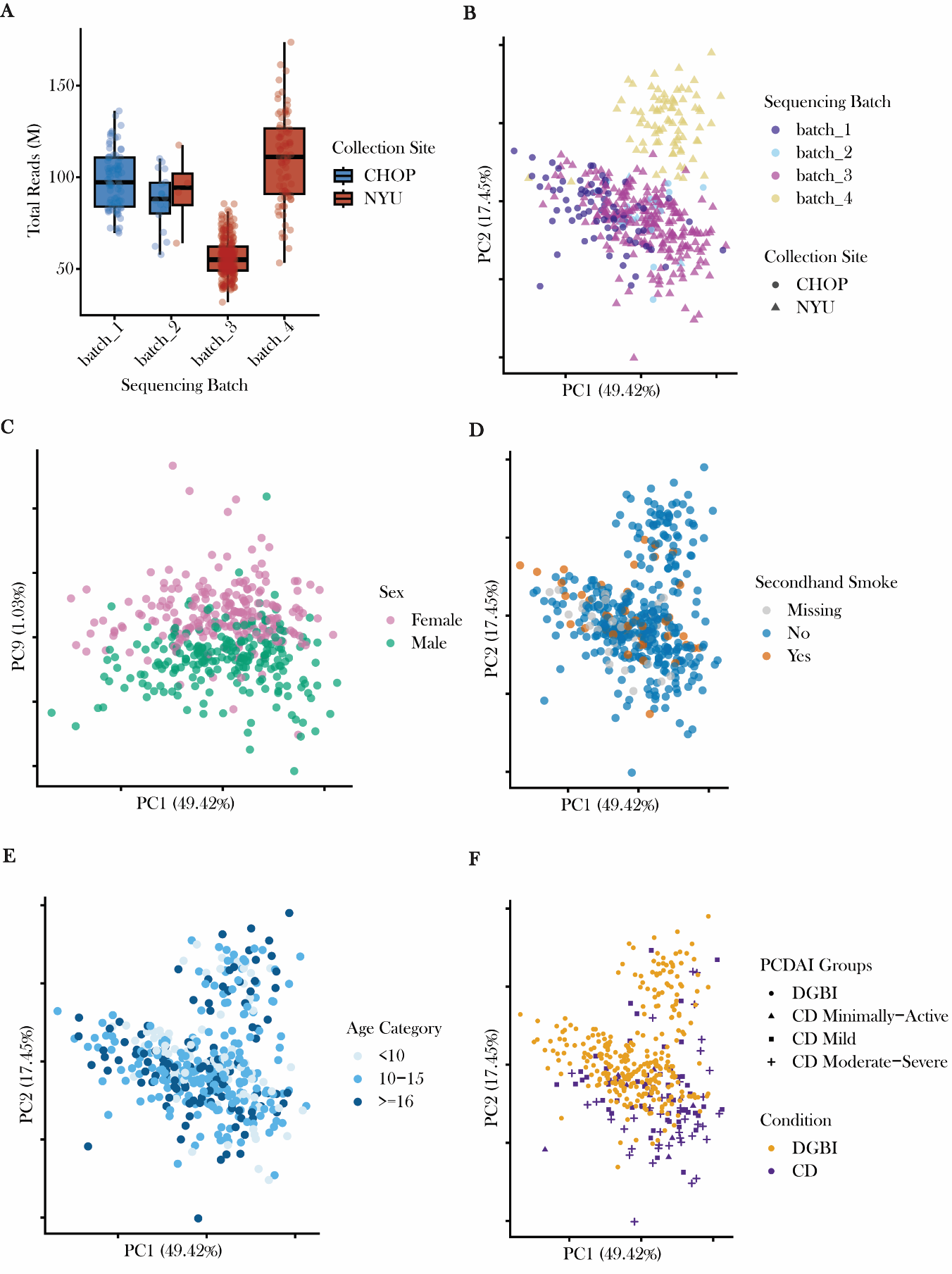

**Supplementary Figure S1. Exploration of whole blood transcriptomes using principal component analysis (PCA) reveals batch effects predominantly explained by sequencing batch**. (**A**) Boxplots of total reads by sequencing batch illustrate varying distributions of sequencing depth that are batch specific. Sample recruitment site was largely confounded by sequencing batches, where samples in batches 3 and 4 belong to NYU (red). (**B**) PCA scatterplot of PC1 and PC2 showed most of the variation was explained by sequencing batch, where batch 4 formed a distinct cluster at the top of the plot, while batches 1-3 clustered together. Sequencing batch is colored, and the shapes are sample recruitment site. (**C**) PCA scatterplot illustrated PC9 varies by sex of sample (males are green, females are pink). Figures (**D**) and (**E**) illustrate that there was little genome-wide variation explained by secondhand smoke exposure and age in our cohort. (**F**) The scatterplot shows that observable variation separated many CD samples from the DGBI samples, predominantly the moderate-severe and mild CD samples.

Alt text: Graphs labeled A to F. A is a boxplot of total read counts per sample by sequencing batch, color by sample recruitment site. B is a principal component scatterplot of PC1 versus principal component 1 versus principal component 2, colored by sequencing batch with point shapes indicating recruitment site. C is a principal component scatterplot showing principal component 9, which varies and is colored by sex. D and E are principal component scatterplots colored by secondhand smoke exposure and age categories, respectively. F is a principal component scatterplot colored by diagnosis and PCDAI activity groups.

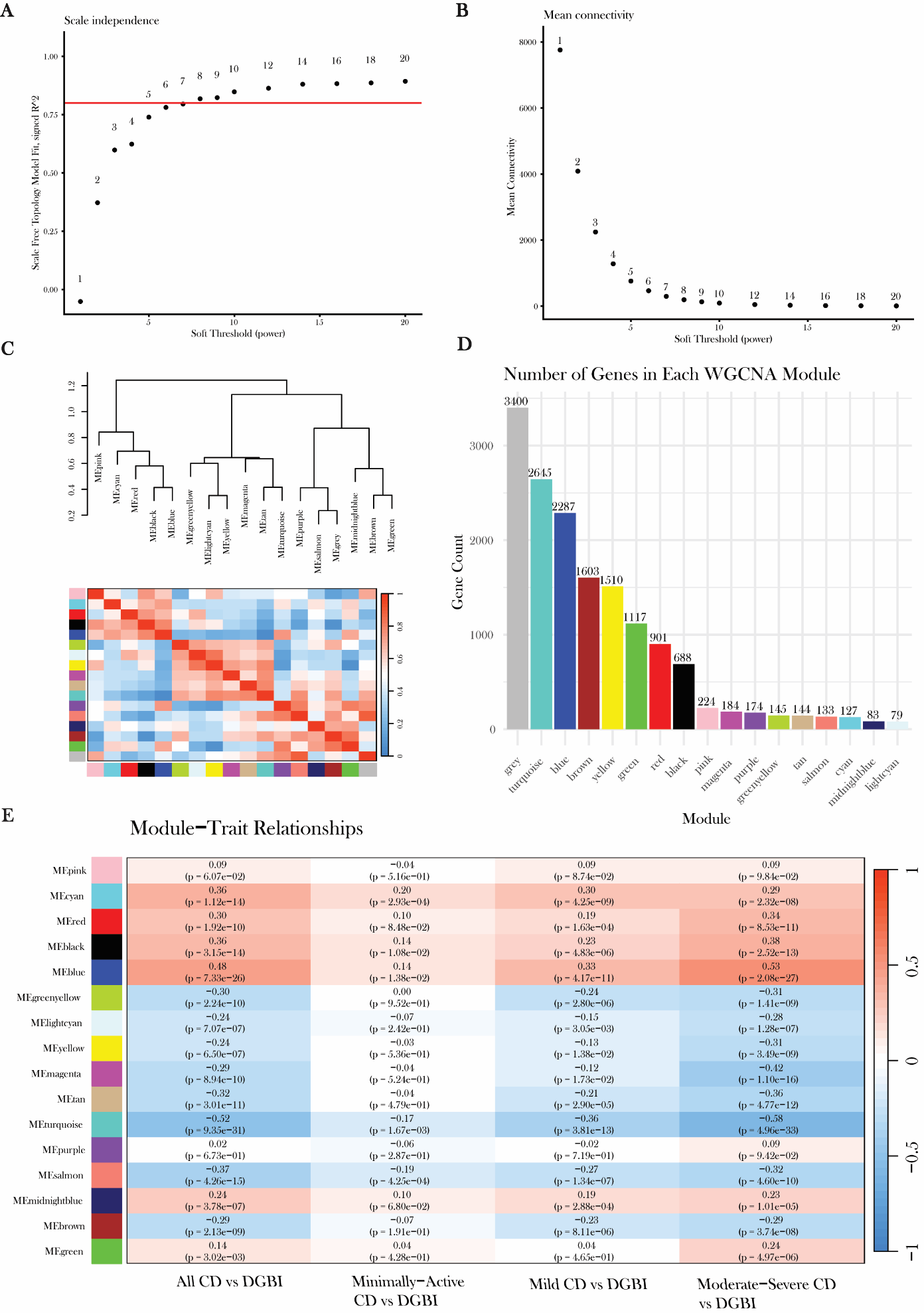

**Supplementary Figure S2. WGCNA identified 16 modules plus the unassigned grey module that varied in association with CD diagnosis and severity.** **(A)** Scale-free topology model fit (signed R²) plotted against candidate soft-thresholding powers. The red horizontal line marks the scale-free fit threshold of R² = 0.80 at soft-threshold power = 8. **(B)** Mean network connectivity at each candidate soft-thresholding power (x-axis). **(C)** Correlation heatmap among module eigengenes (MEs) with hierarchical clustering. Row and column color bars identify the modules, and the color scale represents correlation from -1 (blue) to 1 (red). **(D)** Number of genes assigned to each module, shown as bars colored by module and ordered by size. **(E)** Heatmap of associations between the 16 module eigengenes (rows) and four groups of CD diagnosis and severity: all CD samples versus DGBI, minimally-active CD versus DGBI, mild CD versus DGBI, and moderate-severe CD versus DGBI. The grey module was excluded from downstream analysis. Each cell shows the Pearson correlation with its P value in parentheses. Red indicates a positive association and blue a negative association, on a color scale from −1 to 1.

Alt text: Graphs labeled A to E. A is a scatterplot of signed scale-free topology fit versus soft-threshold power, with a horizontal red line at the 0.80 threshold. B is a scatterplot of mean network connectivity versus soft-threshold power. C is a clustered heatmap with dendrogram of correlations among the module eigengenes, with a color gradient scale showing correlation. E is a heatmap of correlations between the 16 colored module eigengenes and the four CD versus DGBI comparisons, with a color gradient showing the correlation of each group comparison with each module.

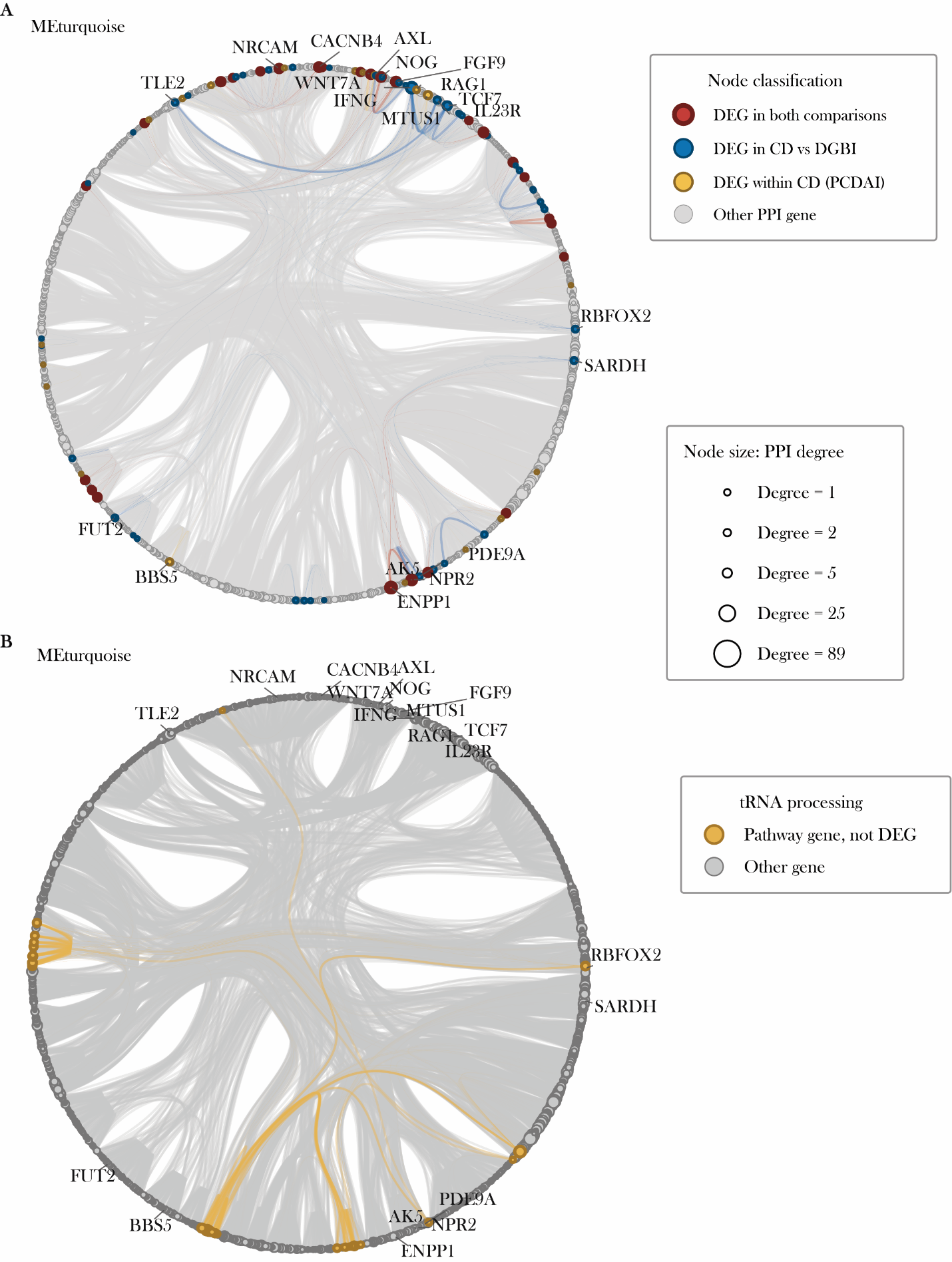

**Supplementary Figure S3. Differentially expressed genes and tRNA processing genes within the turquoise module protein–protein interaction network**. Largest connected component (LCC) of the turquoise module projected onto the STRING protein–protein interaction (PPI) network, shown in the same circular layout in both panels. Nodes represent genes, edges represent PPI interactions with STRING combined score ≥ 500, and node size is proportional to PPI degree (range, 1–89). **(A)** Nodes colored by differential expression status: genes differentially expressed in both CD-vs-DGBI and within-CD by PCDAI (red), in CD-vs-DGBI only (blue), within-CD by PCDAI only (gold), or not a DEG (gray). **(B)** Genes annotated to *tRNA processing* (orange), none of which were differentially expressed. All other gene nodes are shown in gray.

Alt text: Two network diagrams labeled A and B, each showing the same circular layout of the turquoise module’s largest connected component, with nodes grouped by community assignment by a nested stochastic block model. Node size is proportional to the number of connections. A shows the nodes colored by differential expression status, where red are genes significant in both models, blue for CD versus DGBI genes only, gold for PCDAI only, and gray for genes not differentially expressed in either, with edges highlight with the above colors. B highlights the tRNA processing pathway, where pathway genes that are also differentially expressed in either model are colored purple, while nodes colored orange are a pathway gene but not differentially expressed, and gray if neither.

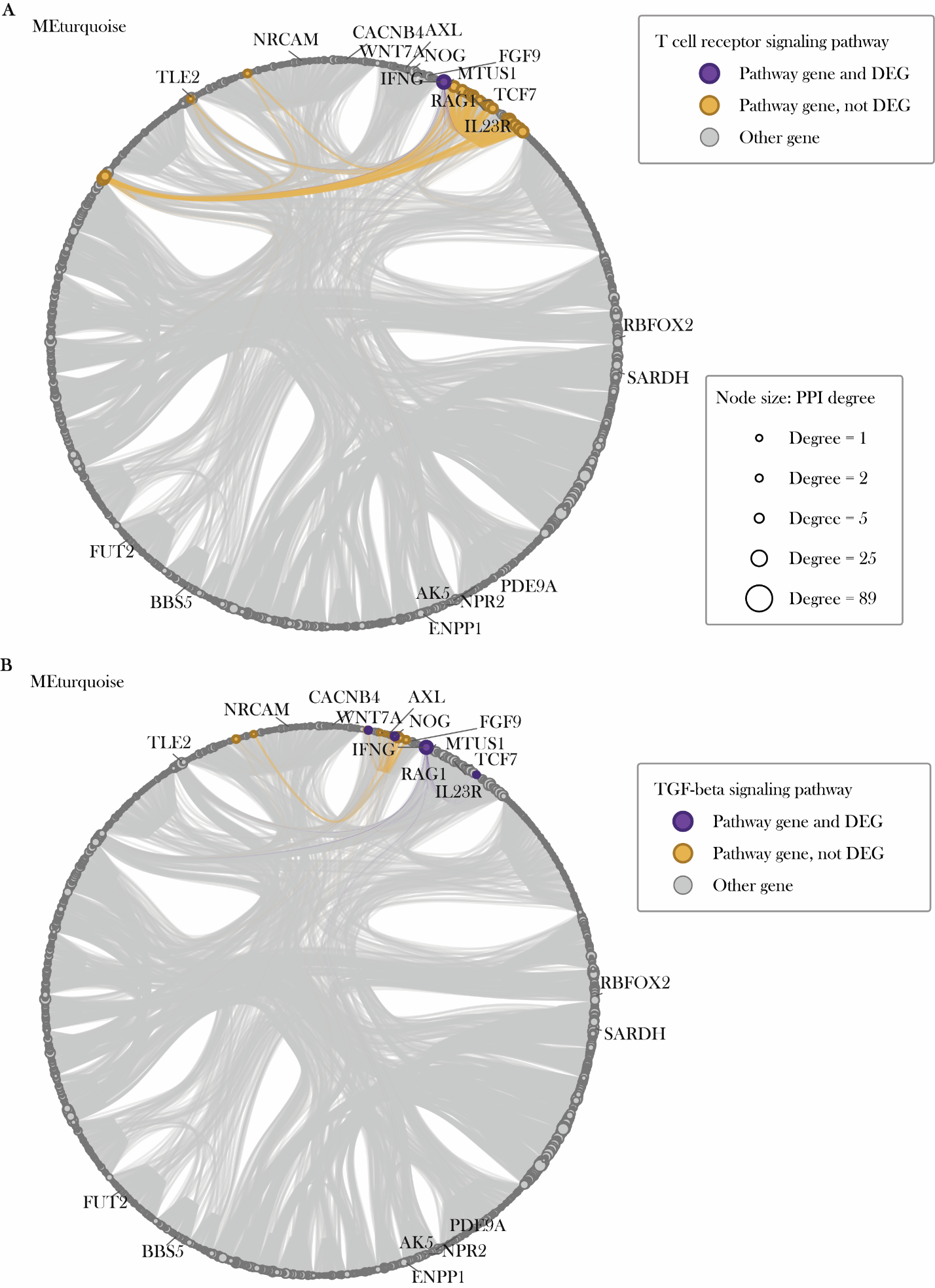

**Supplementary Figure S4. T cell receptor signaling and TGF-beta signaling genes within the turquoise module’s LCC protein–protein interaction network**. PPI network of the turquoise module’s LCC projected from the STRING database, shown in the same circular layout in both panels. Nodes represent genes, gray lines represent PPI edges, and node size is proportional to PPI degree (range, 1–89). **(A)** Genes annotated to the *T cell receptor signaling pathway*, colored by differential expression status: pathway genes that were differentially expressed in either CD-vs-DGBI or within-CD (purple) and pathway genes that were not a DEG (orange). All other genes not in the pathway or not a DEG are shown in gray. **(B)** Genes annotated to the *TGF-beta signaling pathway*, colored as in panel A.

Alt text: Two network diagrams labeled A and B, each showing the same circular layout of the blue module’s largest connected component, with nodes grouped by community assignment by a nested stochastic block model. Node size is proportional to the number of connections. A shows the T-cell receptor signaling pathway and B highlights the TGF-beta signaling pathway, where pathway genes that are also differentially expressed in either model are colored purple, while nodes colored orange are a pathway gene but not differentially expressed, and gray if neither.

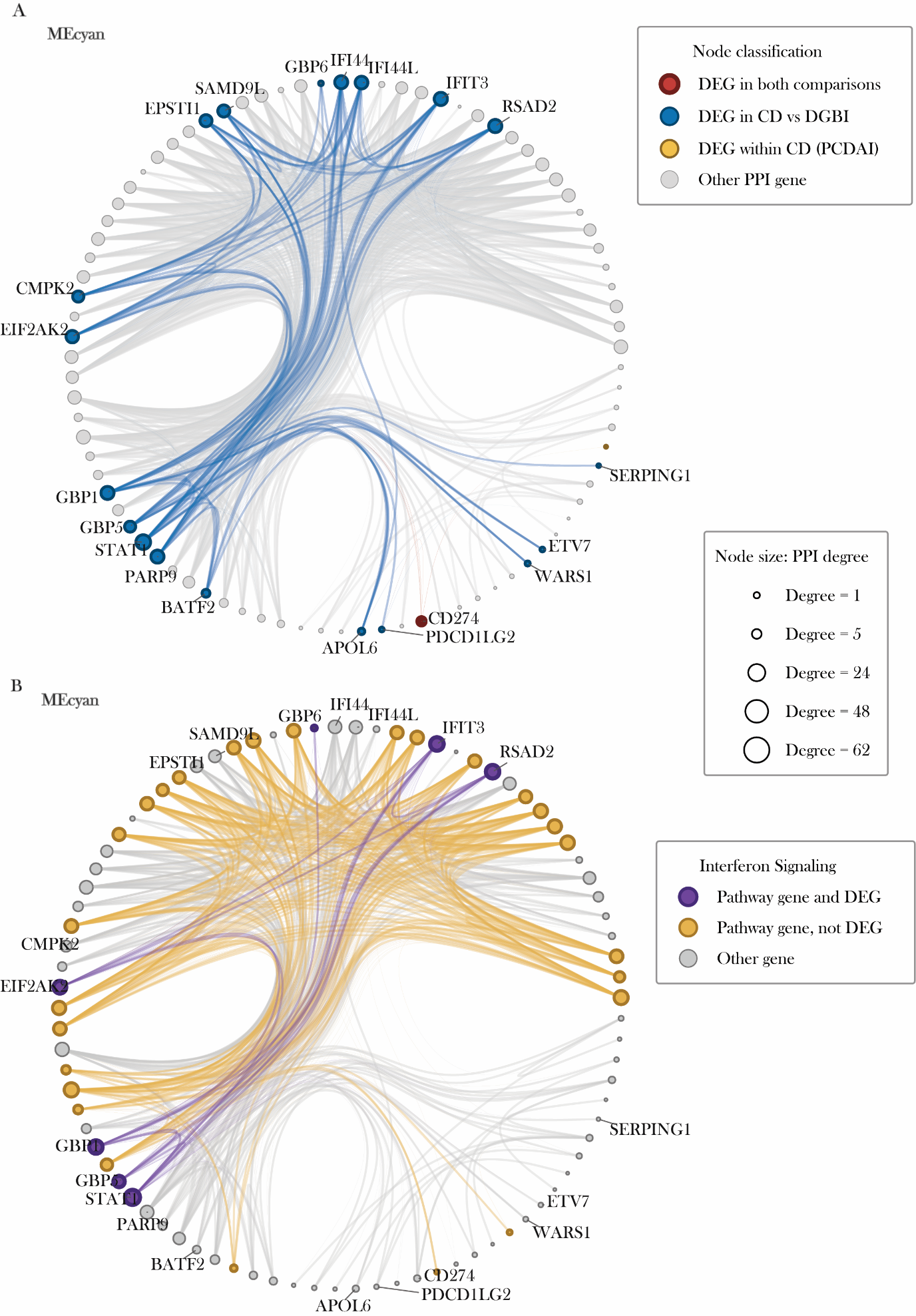

**Supplementary Figure S5. Differentially expressed genes and interferon signaling genes within the cyan module LCC PPI network.** LCC of the cyan module shown in the same circular layout in both panels. Nodes represent genes, gray lines represent PPI edges, and node size is proportional to PPI degree (range, 1–62). **(A)** Nodes colored by differential expression status: genes differentially expressed in both CD-vs-DGBI and within-CD by PCDAI (red), genes differentially expressed in CD-vs-DGBI only (blue), within-CD by PCDAI only (gold), and genes not differentially expressed (gray). **(B)** Genes annotated to *Interferon Signaling*, colored by differential expression status: pathway genes that were differentially expressed in either CD-vs-DGBI or within-CD by PCDAI (purple) and pathway genes that were not DE (orange). All other genes are shown in gray, and edges involving pathway genes are highlighted.

Alt text: Two network diagrams labeled A and B, each showing the same circular layout of the cyan module’s largest connected component, with nodes grouped by community assignment by a nested stochastic block model. Node size is proportional to the number of connections. A shows the nodes colored by differential expression status, where red are genes significant in both models, blue for CD versus DGBI genes only, gold for PCDAI only, and gray for genes not differentially expressed in either, with edges highlight with the above colors. B highlights the interferon signaling pathway, where pathway genes that are also differentially expressed in either model are colored purple, while nodes colored orange are a pathway gene but not differentially expressed, and gray if neither.

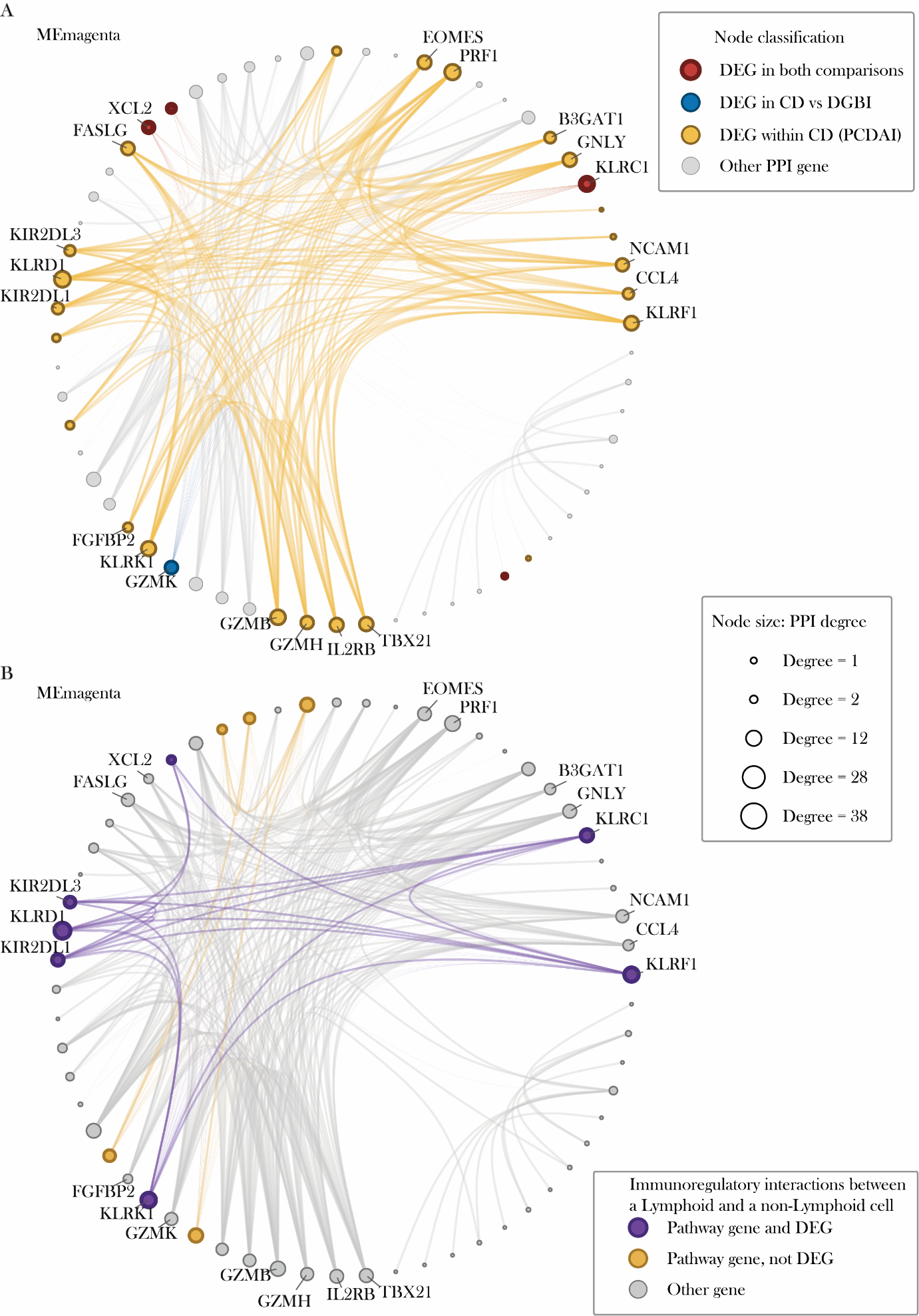

**Supplementary Figure S6. Differentially expressed genes and Immunoregulatory interaction genes within the magenta module protein–protein interaction network.** LCC of the magenta module projected from the STRING, shown in the same circular layout in both panels. Nodes represent genes, gray lines represent PPI edges, and node size is proportional to PPI degree (range, 1–38). **(A)** Nodes colored by differential expression status: genes differentially expressed in both CD-vs-DGBI and within-CD by PCDAI (red), in CD-vs-DGBI only (blue), within-CD by PCDAI only (gold), or in neither comparison (gray). Edges between differentially expressed genes are highlighted. **(B)** Genes annotated to *Immunoregulatory interactions between a lymphoid and a non-lymphoid cell*, colored by differential expression status: pathway genes that were differentially expressed in CD-vs-DGBI or within-CD models (purple) and pathway genes that were not (orange). All other genes are shown in gray, and edges involving pathway genes are highlighted.

Alt text: Two network diagrams labeled A and B, each showing the same circular layout of the magenta module’s largest connected component, with nodes grouped by community assignment by a nested stochastic block model. Node size is proportional to the number of connections. A shows the nodes colored by differential expression status, where red are genes significant in both models, blue for CD versus DGBI genes only, gold for PCDAI only, and gray for genes not differentially expressed in either, with edges highlight with the above colors. B highlights the immunoregulatory interactions between lymphoid and non-lymphoid cells, where pathway genes that are also differentially expressed in either model are colored purple, while nodes colored orange are a pathway gene but not differentially expressed, and gray if neither.

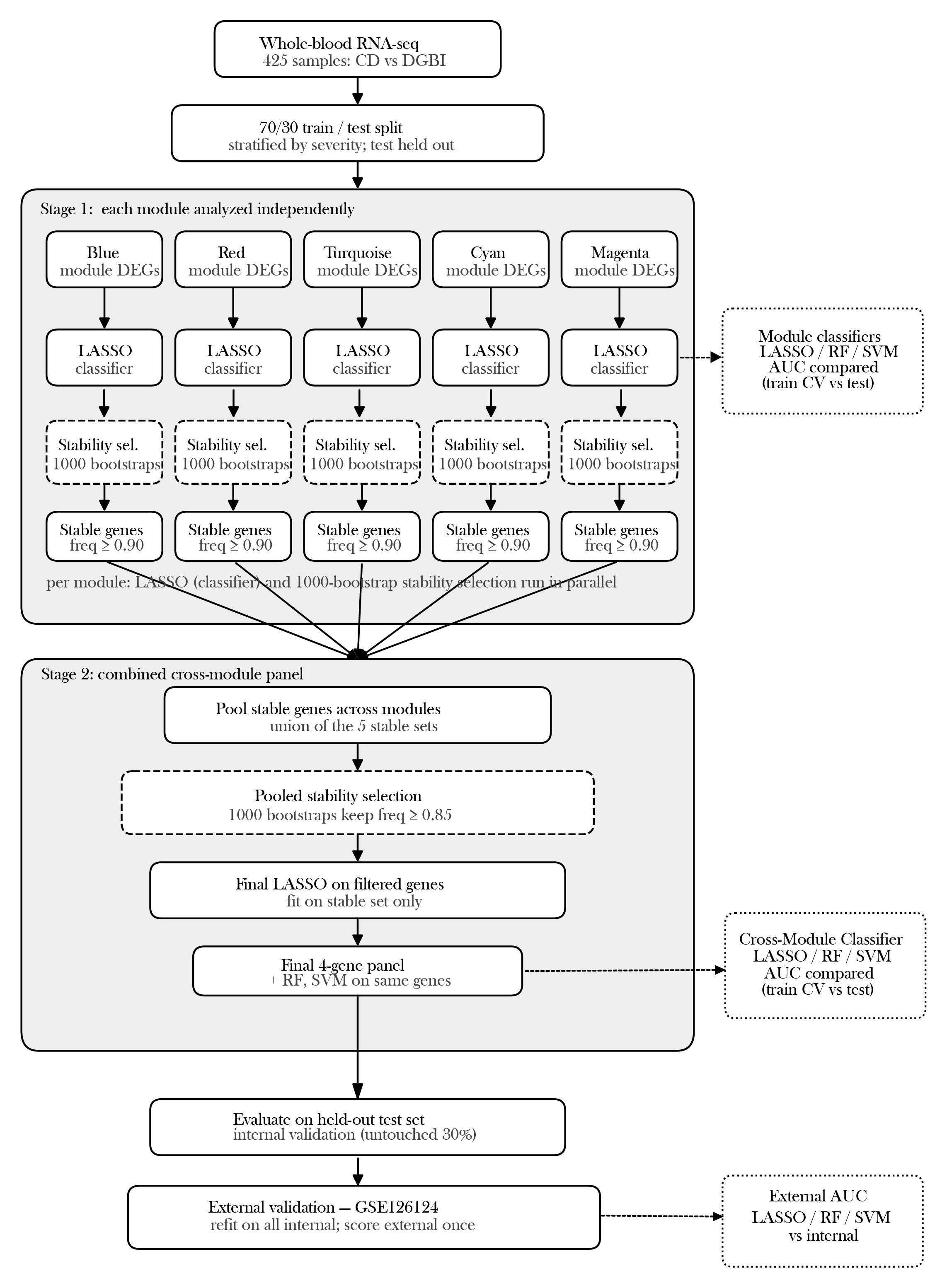
**Supplementary Figure S7. Two-stage classifier development.** Candidate genes (DEGs within each module's STRING largest connected component) were derived from all 425 samples. Samples were then split 70/30 (training n = 297; test n = 128), stratified by diagnosis and CD severity. In Stage 1, each module was analyzed independently. A LASSO fit to the training set defined a locked panel, used to train LASSO, random forest, and SVM classifiers, and 1,000-bootstrap stability selection identified genes selected in >= 90% of resamples. In Stage 2, the stable genes were pooled and re-subjected to a second round of stability selection (>=85%), yielding a compact, cross-module panel. Performance was assessed by cross-validation and on the held-out test set. The panel was then refit on all 425 samples and evaluated once in an external pediatric cohort (GSE126124: 39 CD; 37 symptomatic controls). Dashed boxes, stability selection; dotted boxes, performance outputs.

Alt text: Flowchart of the two-stage classifier pipeline, with boxes and arrows running from candidate gene definition in 425 samples, through a 70/30 training and test split. Stage1 includes the per-module classifiers and stability selection, and Stage 2 pooled stability across modules and produced the 4-gene panel, which was evaluate d by cross-validation in the training, on the held-out test set, and in the external cohort of 39 Crohn’s disease and 37 symptomatic control blood samples.
